# Epigenetic age acceleration links atherogenic dyslipidaemia, inflammageing and frailty with adverse cardiovascular outcomes in older adults (UFO): a lipid-metabolome and epigenetic clock analysis

**DOI:** 10.1101/2025.06.09.25329276

**Authors:** Kwan Hung Ng, Gloria H.W. Lau, Ka Man Yu, Jenny S.W. Lee, Suyi Xie, Leong-Ting Lui, Lihua Huang, Tung Wai AuYeung, Jean Woo, Alice P.S. Kong, Ronald C.W. Ma, Juliana C.N. Chan, Petri Wiklund, Ville-Petteri Mäkinen, Marjo-Riitta Järvelin, Cecilia W. Lo, Stephan Beck, Simone Ecker, Erik Fung

## Abstract

**Background:** Frailty and chronic inflammatory diseases are known risk factors for adverse outcomes and have been associated with epigenetic age acceleration (EAA), a quantitative estimation of biological age based on DNA methylation. We investigated the interrelationships between EAA, atherogenic dyslipidaemia and frailty; determined the best performing epigenetic clock for EAA estimation of atherosclerotic cardiovascular disease (ASCVD) risks; and prospectively analysed the capacity of EAA to predict future adverse outcomes.

**Methods:** A structured cardiogeriatric evaluation including frailty and physical capacity assessment was performed in community-living older adults aged ≥60 years who met predefined eligibility criteria and had no previous history of heart failure. We prioritised the selection of all available frail older adults from our bioresource and used computerised randomisation to select the more abundant robust and pre-frail individuals for relatively balanced analyses among groups. DNA methylation analysis was performed using the Infinium MethylationEPIC platform. Quantitative proton-nuclear magnetic resonance (NMR) was used for targeted metabolomic analysis of serum. Five common epigenetic clocks were compared for associations with frailty, systemic inflammation, lipid-metabolome and prediction of incident adverse outcomes. Clinical outcomes were queried using electronic medical record systems and by interview.

**Findings:** Among 535 older adults, GrimAge 2-estimated EAA (Grim2AA) performed best in classifying and predicting the frailty phenotype. Grim2AA was significantly associated with systemic inflammation indicated by GlycA, increased risk of ASCVD comorbidities, and an atherogenic lipid profile characterised by reduced low-density lipoprotein (LDL) particle size and abnormal triglycerides in lipoproteins. Small LDL particle size but not other lipid/lipoprotein features were also associated with frailty. Of the five epigenetic clocks analysed and benchmarked against the ACC/AHA ASCVD Risk Calculator, Grim2AA was the most strongly associated. For each 10-year increment in Grim2AA, the risk was estimated at 4·92% (p=0·0002). During a median follow-up of 4·68 years, 55 all-cause deaths occurred and 101 individuals experienced cardiovascular hospitalisation. Analysis of cardiovascular hospitalisation revealed marked differences in EAA depending on the cause, and pointed to ASCVD (excluding stroke) as being most common with the highest median Grim2AA at 0·73 years. Kaplan-Meier analysis did not show a difference in the rates of cardiovascular hospitalisation between Grim2AA ≥0 and <0 years (p=0·19). However, the rates of ASCVD hospitalisation (42 of 101) were significantly higher in older adults with the former (p=0·0027).

**Interpretation:** As the most comprehensive targeted analysis of blood lipid-metabolome and DNA methylation-based epigenetic clock to date, this study has linked frailty, inflammageing, atherogenic dyslipidaemia and ASCVD risks that can be collectively indicated by Grim2AA in older adults. Grim2AA is an independent predictor of future ASCVD hospitalisation, and may serve as a potential biomarker for monitoring disease trajectory and target for secondary prevention.

## Introduction

Ageing is dictated by genetic factors and environmental exposures across time that give rise to a range of disorders from immune dysregulation, inflammation (’inflammageing’), cardiometabolic disease, cognitive and physical decline, debilitation to frailty in old age.[James 2024] As leading causes of death and morbidity worldwide, ischaemic heart disease, acute myocardial infarction and stroke are devastating atherosclerotic cardiovascular disease (ASCVD) complications that commonly result from chronic dyslipidaemia and maladapted low-grade systemic inflammation. Conventionally, atherogenic dyslipidaemia is characterised by increased blood concentration of proinflammatory small dense low-density lipoprotein (LDL) particles and triglycerides, and reduced high-density lipoprotein (HDL) cholesterol level. However, the conceptual definition has evolved over time to include other triglyceride-rich lipoproteins (TRLs) and apolipoproteins.[Mach 2020] With advances in high-field proton (^1^H)-nuclear magnetic resonance (NMR) for in vitro diagnostic research during the last decade that can substratify and analyse blood lipids and lipoproteins in a rapid (i.e. turnaround time on the order of minutes), reproducible (typically, percent coefficient of variation of between 1% and 5%) and non-destructible (repeat measurements possible) manner,[Soininen 2015] the technology has opened up new avenues for re-examining and redefining the concept of atherogenic dyslipidaemia.

ASCVD risk prediction remains challenging due in part to the long disease latency, ethnic-dependent variability, unpredictable trajectory, and variable activity of the disease. Available studies have shown that traditional variables (e.g. total cholesterol and LDL-cholesterol concentrations, blood pressure, diabetes status) while clinically practical in multifactorial risk assessment when benchmarked against a population-wide reference dataset,[Lloyd-Jones 2019; Visseren 2021; Hippisley-Cox 2017; SCORE-OP 2021] lack personalised precision and granularity for higher performance. Indeed, Dalton et al. pointed out the inadequacy and failure of traditional risk factors to predict cardiovascular events in a study of over 25,000 patients aged ≥65 years at study baseline.[Dalton 2020] The urgent need to more deeply characterise and predict risks in ASCVD and to treat it earlier with precision has been pointed out by Makover et al.[Makover 2022]

One of the shortcomings of the available ASCVD risk prediction tools is the limited data on older adults, particularly the old-old (age 75–84 y) and the oldest old (age ≥85 y). For instance, among the most widely used ACC/AHA ASCVD Risk Calculator, risk assessment is only possible in individuals within the age range of 40– 79 years. Similarly, the original ESC HeartScore^®^ had a limited age range of 45–64 years,[Conroy 2003] prompting subsequent development of SCORE (age range 40–69 y) and SCORE-OP (age range of 70–89 y).[SCORE-OP 2021]

Age remains one of the most powerful determinants of ASCVD risk and frailty. With increasing age, the divergence between chronological and biological age in each person can impact the accuracy of disease risk estimates. First reported over a decade ago, Horvath et al. used 353 differentially methylated positions (DMPs) in genomic DNA to serve as indicator of chronological age.[Horvath 2013] Subsequent to the first-generation epigenetic clocks of Horvath’s [Horvath 2013] and Hannum’s [Hannum 2013] that approximated chronological age, the second-generation PhenoAge [Levine, 2018] and GrimAge[Lu 2019] were adjusted for chronological age and predicted lifespan and ageing-related adverse outcomes. Extending from GrimAge, GrimAge2 further included additional DMPs as surrogate markers of C-reactive protein (CRP) and haemoglobin A1c levels,[Lu 2022] reflecting the importance of inflammageing and cardiometabolic disease in the life course and ageing. Importantly, recent studies have shown that epigenetic age acceleration (EAA), defined by the residual from regressing epigenetic age on chronological age,[Ecker 2019] can inform about diseases or disease status (e.g. cancer, diabetes), environmental exposures (e.g. smoking), and adverse cardiometabolic risks and adverse outcomes.[Marioni 2015; Perna 2016; Horvath 2018; Ecker 2019; Cardenas 2022; Engelbrecht 2022; Chervova 2023] We reason that the combined use of epigenetic clock and quantitative ^1^H-NMR to profile the circulating blood lipid-metabolome and low-grade systemic inflammation (e.g. glycosylated acetyls (GlycA))[Ojanen 2021; Fung 2023]) improve our understanding of ASCVD risks in the old-old and oldest old beyond what is currently available, and provide predictors of adverse outcomes. In this study of community-living older adults, we aim to 1) characterise the abnormalities in the circulating blood lipid-metabolome that define dyslipidaemia associated with ASCVD comorbidities, inflammageing, frailty, and EAA; and 2) prospectively identify predictors of incident adverse cardiovascular outcomes.

## Methods

### Study participants

The <u>U</u>ndiagnosed heart Failure in frail <u>O</u>lder individuals (UFO) cohort is an ongoing prospective, longitudinal observational study established to characterise the epidemiological factors that were associated with frailty and cardiac dysfunction, heart failure, cardiovascular complications, and other adverse clinical outcomes in community-living older adults across all 18 Districts of Hong Kong Special Administrative Region of China.[Fung 2018] Study participants aged ≥60 years were recruited into the study if they had no prior history of heart failure, met the predefined eligibility criteria, and were able to provide written informed consent. Excluded were individuals who had complex congenital heart disease with or without surgical correction, an implantable cardiac pacemaker or cardioverter defibrillator, ventricular assist device, and heart transplant. In this analysis, we first included all frail individuals in the UFO cohort assessed by the FRAIL scale, then computationally selected at random pre-frail and robust individuals in the remaining cohort to achieve an approximate equal ratio across all three frailty groups to maximise statistical power. All study participants provided written informed consent. The study has been approved by the institutional clinical research ethics committee and adheres to the Declaration of Helsinki.

### Frailty assessments

The FRAIL scale comprises five components (<u>F</u>atigue, <u>R</u>esistance, <u>A</u>mbulation, Illness and <u>L</u>oss of weight)[Abellan van Kan 2008; Morley 2012] that were assessed at the time of recruitment.[Fung 2018] Each positive response was scored 1 point out of a total maximum of 5 points, with ≥3 indicating frail, 1–2 pre-frail, and 0 robust status.[Woo 2019] The disease inventory-based, digital electronic version of the frailty index (eFI) of Clegg et al. based on the cumulative deficit model of Rockwood and Mitniski was also applied and the results were shown where appropriate;[Clegg 2016] briefly, eFI is calculated as a ratio of the number of deficits present to the total number deficits assessed, and categorises frailty into four classes: robust, 0–0·12; mildly frail, 0·12– 0·24; moderately frail, 0·24–0·36; and severely frail, >0·36.[Clegg 2016; Fung 2021] Physical activity and fitness were assessed using 6-minute walk distance (6MWD), gait speed, and handgrip strength by dynamometry.[Woo 2019; Yang 2020; Fung 2021] All frailty assessments were done at the time of recruitment into the study.

### Definition of atherosclerotic cardiovascular disease (ASCVD) and comorbidities

A past history of angina pectoris, ischaemic heart disease (IHD), myocardial infarction, atrial fibrillation, stroke, and other comorbidities including hypertension (defined by blood pressure ≥140/≥90 mmHg),[WHO 2023] diabetes mellitus (defined according to the World Health Organization), [WHO 2020] and chronic kidney disease (CKD) [Jin 2024] were diagnosed in accordance with international guidelines and standards, and according to electronic health records and self-reported questionnaires. Estimated glomerular filtration rate (eGFR) was calculated using the CKD-Epidemiology Collaboration (CKD-EPI) equation. Stroke, IHD, and myocardial infarction were analysed as separate variables, or as a composite ASCVD variable in regression models.

### DNA methylation analysis

Peripheral venous blood was collected from study participants in serum-separating clot activator tubes (Vacuette, Greiner Bio-One GmbH, Austria) and centrifuged within 2–3 hours. Following removal of blood serum, the cell fraction was stored at −80°C until DNA isolation using the Gentra Puregene Blood Kit (Qiagen, Germany). Quality control and quantification of DNA was performed before transport of samples to Novogene (Hong Kong SAR) for interrogation of DNA methylation using the Infinium MethylationEPIC v1.0 BeadChip (Illumina, Inc., San Diego, California, USA). Raw data were imported and analysed using the R package, ChAMP.[Tian 2017] One individual’s sample failed quality control and filtering due to high CpG fraction, while another individual was subsequently found to meet the UFO study’s exclusion criteria and was removed, resulting in a data set of 535 individuals that qualified for subsequent analysis. The filtering process further removed probes with high detection p value, low bead count, or non-CpG nature. Two data normalization methods, NOOB and BMIQ, were performed sequentially on the filtered data.[Welsh 2023]

### Calculation of epigenetic age and epigenetic age acceleration (EAA)

The normalised dataset was submitted to the online DNA Methylation Age Calculator (https://dnamage.clockfoundation.org/) to perform calculations for the respective epigenetic clocks. Five epigenetic clocks used in this study were the Hannum (2013),[Hannum 2013] Horvath (2013),[Horvath 2013] PhenoAge (2018),[Levine 2018] GrimAge (2019)[Lu 2019] and GrimAge2 (2022)[Lu 2022] epigenetic clocks. EAA was defined as the residual values from the regression plot of epigenetic age against chronological age,[Ecker 2019] and was calculated for each epigenetic clock corresponding to HannumAA, HorvathAA, PhenoAA, GrimAA and Grim2AA, respectively.

### Proton-nuclear magnetic resonance (^1^H-NMR) spectroscopy

Blood serum specimens were stored at or below −80°C until transport on dry ice via overnight express courier to Nightingale Health Ltd (Helsinki, Finland) for ^1^H-NMR metabolomic analysis. Blood serum metabolites, amino acids, fatty acids, lipids, lipoproteins, glycoprotein acetylation, low-molecular weight protein and other small biochemical substances were quantified using Bruker AVANCE III spectroscopy in a proprietary pipeline, as described previously.[Soininen 2015] Briefly, prior to analysis, thawed serum were mixed 1:1 with a sodium phosphate buffer (75 mmol/L Na_2_HPO_4_ in 80%/20% H_2_O/D_2_O at pH 7.4 including 0.08% sodium 3- (trimethylsilyl)propionate-2,2,3,3-d_4_ and 0.04% NaN_3_).[Soininen 2015] Samples were handled by an automated temperature-controlled (6°C) liquid handler, and loaded by a robotic sample changer. Initial data processing included Fourier transformations to NMR spectra and automatic phasing, followed by subsequent automated spectral processing and quality control. Regression modelling was performed to generate the quantified molecular data.[Soininen 2015]

### ASCVD risk estimation and modelling

ASCVD risk estimates were calculated based risk equations from the ACC/AHA (2013) ASCVD Risk Calculator published previously.[Lloyd-Jones 2019; Lloyd-Jones 2019; Medina-Inojosa 2023] Risk estimator parameters originally developed for white men and women were used in our analysis. As systolic blood pressure measurements were not available in approximately ∼60% of the study participants in the early phased of participant recruitment, multiple imputation was performed for the missing data using Amelia v1.8, an R software package.[Honaker 2011] 1000 datasets with imputed blood pressure data and the calculated ASCVD risk were generated for statistical analysis. Multiple regression was performed to study the association between EAA with/without lipidome-metabolome biomarkers and ASCVD risks on each imputed dataset. Results were summarised in the distribution of beta values and p values of the dependent variables.

### Statistical analysis

The Cochran-Armitage test was used to compare categorical variables across FRAIL classes. The Jonckheere-Terpstra trend test was used for testing of non-parametric continuous variables. Spearmen correlation coefficients between each epigenetic clock model and chronological age were calculated and summarised in the correlation matrices. Multiple linear regression was used to determine the effects of EAA on frailty assessments, and the shortlisted lipid-metabolomic biomarkers. Logistic regression was performed to analyse the effect size of EAA on binary comorbidities and disease-associated variables, and expressed as adjusted odds ratio (OR). All models were adjusted for age, sex, smoking status, and other variables as indicated. To address false discovery rate (FDR) associated with multiple testing in lipid-metabolome and EAA analyses, the Benjamini-Hochberg procedure was used. Cox proportional hazards models were developed by regressing on dependent variables of incident outcomes, including all-cause death and hospitalisation, and expressed as adjusted hazard ratios (HRs). Cardiovascualar hospitalisation was defined as hospitalisation due to a cardiovascular cause. ASCVD hospitalisation was defined as hospitalisation due to acute coronary syndrome (ST-elevation and non-ST-elevation myocardial infarction and unstable angina), angina pectoris, ischaemic heart disease requiring percutaneous coronary intervention, peripheral vascular disease including acute limb ischaemia and ruptured or unruptured aortic aneurysm requiring hospital admission and intervention(s). Kaplan-Meier analysis was used to plot cumulative events and determine statistical significance between EAA Grim2AA <0 and ≥0 years. A p value of <0·05 was considered statistically significant.

## Results

### 1. Characteristics of study participants

Among 535 community-living older adults in the cohort, the median ages for the robust (n=186/535, 34·8%), pre-frail (n=180/535, 33·6%) and frail (n=169/535, 31·6%) groups were 71 y (IQR 66–83, range 60–92), 80 y (IQR 70–85, range 60–94) and 82 y (IQR 75–86, range 63–99), respectively, corresponding to median eFI values of 0·06 [IQR 0·03–0·08], 0·14 [IQR 0·08–0·19] and 0·22 [IQR 0·17–0·28], respectively (Jonckheere-Terpstra test, p<0·0001). Based on a median eFI of 0·22, the majority of participants in the frail group were mild-to-moderately frail.[Clegg 2016; Fung 2021]

Overall, increased frailty was associated with older age (p<0·0001), female preponderance (p<0·0001), worse renal function by eGFR (p<0·0001), and higher prevalence of cardiovascular, renal and metabolic comorbidities (hypertension, p<0·0001; diabetes mellitus, p<0·0001; angina pectoris, p<0·0001; IHD, p<0·0001; myocardial infarction, p<0·05; stroke, p<0·0001; CKD, p<0·0001) (**Table 1**). Incremental frailty was associated with decreased 6MWD, gait speed and hand grip strength (all p<0·0001) (**Table 1**).

**Table 1.** Baseline characteristics of participants in the UFO study on epigenetic age, frailty, inflammageing and ASCVD. See Appendix p 2 for raw p values.

| Variable | All | Robust<br>(FRAIL 0) | Pre-frail<br>(FRAIL 1–2) | Frail<br>(FRAIL ≥3) | p value |
| --- | --- | --- | --- | --- | --- |
| N | 535 | 186 | 180 | 169 | - |
| Median age (IQR), y | 78 (69–85) | 71 (66–83) | 80 (70–85) | 82 (75–86) | <0.0001 |
| Median age (range, min–max), y | 78 (60–99) | 71 (60–92) | 80 (60–94) | 82 (63–99) | - |
| Sex (% male), *n* | 177 (33.1%) | 90 (48.4%) | 53 (29.4%) | 34 (20.1%) | <0.0001 |
| Body mass index, *kg/m*<sup>2</sup> | 24.03 (21.88–26.62) | 23.46 (21.76–25.42) | 24.52 (21.95–27.16) | 24.35 (21.91–27.27) | <0.05 |
| Smoking history (%) | 110 (20.6%) | 49 (26.3%) | 33 (18.3%) | 28 (16.6%) | <0.05 |
| Past alcohol use (%) | 116 (21.7%) | 50 (26.9%) | 40 (22.2%) | 26 (15.4%) | <0.01 |
| eGFR*, *mL/min/1.73 m*<sup>2</sup> | 79.23 (63.24–89.71) | 85.51 (74.50–94.12) | 75.78 (61.07–87.84) | 71.13 (51.01–85.52) | <0.0001 |
| **Frailty measurements** |  |  |  |  |  |
| eFI<sup>†</sup> | 0.11 (0.06–0.19) | 0.06 (0.03–0.08) | 0.14 (0.08–0.19) | 0.22 (0.17–0.28) | <0.0001 |
| 6-minute walk distance, *m* | 326 (259–386) | 380 (340–426) | 320 (248–380) | 258 (189–308) | <0.0001 |
| Gait speed, *m/s* | 0.92 (0.71–1.13) | 1.17 (1.04–1.30) | 0.86 (0.70–1.02) | 0.73 (0.58–0.87) | <0.0001 |
| Handgrip strength, *kg* | 18 (14–23) | 22 (18–27) | 17 (14–22) | 15 (13–18) | <0.0001 |
| **Comorbidities** |  |  |  |  |  |
| Hypertension | 315 (58.9%) | 72 (38.7%) | 116 (64.4%) | 127 (75.1%) | <0.0001 |
| Diabetes mellitus | 117 (21.9%) | 14 (7.5%) | 44 (24.4%) | 59 (34.9%) | <0.0001 |
| Angina pectoris | 49 (9.2%) | 3 (1.6%) | 12 (6.7%) | 34 (20.1%) | <0.0001 |
| Atrial fibrillation | 18 (3.4%) | 6 (3.2%) | 7 (3.9%) | 8 (4.7%) | 0.10 |
| ASCVD<sup>‡</sup> (IHD, MI and/or stroke) | 91 (17.0%) | 9 (4.8%) | 31 (17.2%) | 51 (30.2%) | <0.0001 |
| IHD | 46 (8.6%) | 6 (3.2%) | 11 (6.1%) | 29 (17.2%) | <0.0001 |
| MI | 10 (1.9%) | 2 (1.1%) | 1 (0.6%) | 7 (4.1%) | <0.05 |
| Stroke | 53 (9.9%) | 2 (1.1%) | 20 (11.1%) | 31 (18.3%) | <0.0001 |
| Chronic kidney disease | 31 (5.8%) | 2 (1.1%) | 7 (3.9%) | 22 (13%) | <0.0001 |

ASCVD, atherosclerotic cardiovascular disease; eFI, electronic frailty index; eGFR, estimated glomerular filtration rate; IHD, ischemic heart disease; IQR, interquartile range; MI, myocardial infarction.
For categorical and continuous variables, the Cochran-Armitage test and the Jonckheere-Terpstra trend test were used, respectively. Raw p values are shown in appendix p 2.
\*Calculated using the Chronic Kidney Disease Epidemiology Collaboration (CKD-EPI) formula
<sup>†</sup>Calculated by the method of Clegg A et al.<sup>[REF]</sup> using retrievable data from 438 individuals.
<sup>‡</sup>Composite variable of stroke, IHD and/or MI.

### 2. Frailty, epigenetic clock and epigenetic age acceleration (EAA)

Increased epigenetic age was associated with increased frailty as assessed by the FRAIL scale (p=8·58 x 10^−20^ to 1·17 x 10^−8^) or eFI (p=1·80 x 10^−35^ to 6·34 x 10^−19^) (**Table 2 & Appendix p 3**). All five epigenetic clocks — Horvath, Hannum, PhenoAge, GrimAge, and GrimAge2 — examined in this study were positively correlated with chronological age (Spearman’s π=0·58–0·82, all adjusted p<0·0001) (**Figure 1**). Among the clocks, GrimAge and GrimAge2 were most strongly correlated (π=0·77–0·82) (**Figure 1**) and showed no sex differentiated effects, irrespective of FRAIL or eFI (male, π=0·79–0·85 and female, π=0·79–0·86; **Appendix pp 4–5**).

**Figure 1.**
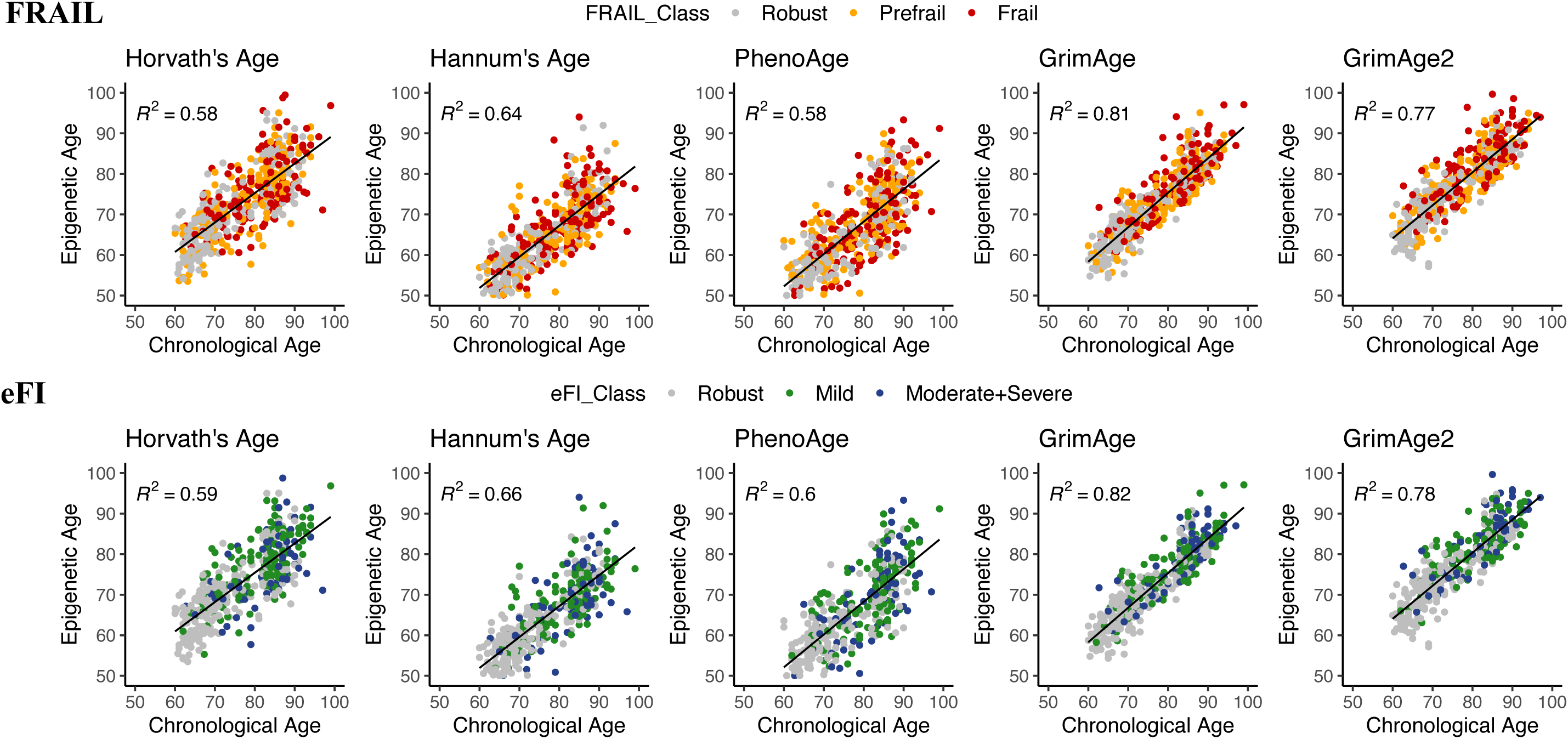
Spearman correlation of epigenetic age with chronological age. FRAIL (robust, *grey*; pre-frail, *orange*; frail, *red*) and eFI (non-frail/robust, *grey*; mildly frail, *green*; moderate-to-severely frail, *blue*) classification of frailty classes are shown. All p values <0·0001. See Appendix pp 4 and 5 for sex-stratified analysis of male and female study participants, respectively.

**Table 2.** Association between frailty and epigenetic age or epigenetic age acceleration (EAA).

| Characteristic | All | Robust<br>(FRAIL 0) | Pre-frail<br>(FRAIL 1–2) | Frail<br>(FRAIL ≥ 3) | p value |
| --- | --- | --- | --- | --- | --- |
| N | 535 (100%) | 186 (34.8%) | 180 (33.6%) | 169 (31.6%) | - |
| **Epigenetic clock** |  |  |  |  |  |
| Horvath's Age (2013), y | 72.79 (66.96–79.24) | 69.71 (64.79–75.92) | 74.20 (67.70–79.92) | 74.52 (70.21–79.71) | <0.0001 |
| Hannum's Age (2013), y | 65.03 (54.97–68.40) | 60.30 (54.97–68.40) | 65.42 (58.81–72.32) | 67.29 (62.00–72.45) | <0.0001 |
| PhenoAge (2018), y | 65.40 (57.73–72.81) | 59.90 (54.28–69.42) | 67.20 (60.05–73.52) | 68.27 (62.25–74.43) | <0.0001 |
| GrimAge (2019), y | 73.74 (66.27–79.44) | 68.20 (62.77–77.69) | 74.50 (67.60–79.46) | 76.50 (70.81–81.37) | <0.0001 |
| GrimAge2 (2022), y | 79.23 (71.57–84.45) | 73.63 (68.08–82.12) | 79.44 (73.39–84.19) | 81.82 (76.14–86.62) | <0.0001 |
| **Epigenetic age acceleration** |  |  |  |  |  |
| Horvath's AA, $\beta$ | 0.04 (-4.08 to 3.77) | -0.09 (-3.82 to 3.71) | 0.09 (-3.78 to 3.85) | -0.29 (-4.58 to 3.65) | 0.44 |
| Hannum's AA, $\beta$ | -0.45 (-3.58 to 2.97) | -0.50 (-3.15 to 2.69) | -0.29 (-3.23 to 2.93) | -0.52 (-3.99 to 3.17) | 0.98 |
| PhenoAA, $\beta$ | -0.24 (-4.02 to 3.70) | 1.01 (-4.28 to 2.47) | 0.40 (-3.35 to 4.62) | -0.24 (-4.27 to 4.47) | 0.24 |
| GrimAA, $\beta$ | -0.42 (-2.56 to 2.12) | -0.52 (-2.46 to 1.82) | -0.52 (-2.65 to 1.71) | -0.35 (-2.65 to 2.56) | 0.49 |
| Grim2AA, $\beta$ | -0.51 (-2.69 to 2.34) | -0.83 (-3.00 to 1.72) | -0.40 (-2.92 to 2.21) | 0.02 (-2.46 to 3.52) | <0.05 |

\*The Jonckheere-Terpstra trend test were used to calculate p values for continuous variables

Association analysis of EAA with FRAIL, eFI and physical frailty measures including 6-minute walk distance, gait speed, and handgrip strength identified GrimAA (with FRAIL, adjusted β 0·096 [95% CI, 0·064– 0·128], FDR-adjusted p=4·71×10^−8^; with eFI, adjusted β 0·007 [95% CI, 0·004–0·009], FDR-adjusted p=6·6 x 10^−7^) and Grim2AA (with FRAIL, adjusted β 0·089 [95% CI, 0·063–0·116], FDR-adjusted p=1·38 x 10^−9^; with eFI, adjusted β 0·006 [95% CI, 0·004–0·008], FDR-adjusted p=4.43 x 10^−8^) as the most strongly associated epigenetic clocks for estimation of EAA and frailty (Table 3 & Appendix p 6); whereas HannumAA, PhenoAA and HorvathAA showed weak or no association (Table 3).

**Table 3.** Multiple regression of epigenetic age acceleration (EAA) on FRAIL, eFI, physical fitness measures, and the composite NMR marker of systemic inflammation, GlycA.

| Dependent variables | Horvath's AA |  | Hannum's AA |  | PhenoAA |  | GrimAA |  | Grim2AA |  |
| --- | --- | --- | --- | --- | --- | --- | --- | --- | --- | --- |
| | $\beta$ (95% CI) | FDR-adjusted p value | $\beta$ (95% CI) | FDR-adjusted p value | $\beta$ (95% CI) | FDR-adjusted p value | $\beta$ (95% CI) | FDR-adjusted p value | $\beta$ (95% CI) | FDR-adjusted p value |
| FRAIL | 0.007 (-0.013 to 0.026) | 0.52 | 0.02 (-0.001 to 0.041) | 0.09 | 0.02 (0.004–0.036) | <0.05 | 0.096 (0.064–0.128) | <0.0001 | 0.089 (0.063–0.116) | <0.0001 |
| eFI | 0.001 (-0.001 to 0.002) | 0.42 | 0.002 (0–0.003) | <0.05 | 0.001 (0–0.002) | 0.06 | 0.007 (0.004–0.009) | <0.0001 | 0.006 (0.004–0.008) | <0.0001 |
| 6-min walk distance | -1.177 (-2.538 to 0.184) | 0.12 | -2.318 (-3.846 to -0.79) | <0.01 | -1.414 (-2.53 to -0.298) | <0.05 | -5.336 (-7.633 to -3.039) | <0.0001 | -5.022 (-6.92 to -3.125) | <0.0001 |
| Gait speed | -0.006 (-0.01 to -0.001) | <0.05 | -0.009 (-0.014 to -0.004) | <0.001 | -0.004 (-0.007 to 0) | 0.06 | -0.017 (-0.024 to -0.01) | <0.0001 | -0.014 (-0.02 to -0.008) | <0.0001 |
| Handgrip strength | 0.035 (-0.049 to 0.118) | 0.45 | -0.078 (-0.174 to 0.017) | 0.13 | -0.01 (-0.08 to 0.06) | 0.77 | -0.168 (-0.309 to -0.027) | <0.05 | -0.155 (-0.272 to -0.038) | <0.05 |
| GlycA | -0.002 (-0.005 to 0.002) | 0.43 | 0.003 (-0.001 to 0.007) | 0.21 | 0.005 (0.002–0.008) | <0.01 | 0.019 (0.013–0.024) | <0.0001 | 0.017 (0.012–0.022) | <0.0001 |

$\beta$ value indicates the predicted change in the dependent variables per 1-year increase in EAA. CI, confidence interval; eFI, electronic frailty index; FDR, false discovery rate; GlycA, glycosylated acetyls. Raw p values are shown in Appendix p 6.

### 3. EAA and cardiometabolic comorbidities

Using logistic regression to model per one-year increase in EAA on common cardiovascular and non-cardiovascular diseases (comorbidities) as dependent variables (**Table 4**), it was determined that increased GrimAA or Grim2AA was associated with conditions that are known to increase the risks for ASCVD including diabetes mellitus (GrimAA, OR 1·017 [95% CI, 1·006–1·028], p=0·0146; Grim2AA, OR 1·018 [1·009–1·027], p=0·0028), angina pectoris (GrimAA, OR 1·011 [1·004–1·019], p=0·0169; Grim2AA, OR 1·001 [1·004– 1·017], p=0·0105), stroke (GrimAA, OR 1·01 [1·002–1·018], p=0·0373; Grim2AA, OR 1·009 [1·003–1·016], p=0·0225), IHD (GrimAA, OR 1·01 [1·003–1·017], p=0·0225; Grim2AA, OR 1·001 [1·004–1·016], p=0·0101), and myocardial infarction (GrimAA, OR 1·05 [1·002–1·009], p=0·022; Grim2AA, OR 1·004 [1·001–1·007], p=0·0225). The composite ASCVD variable of stroke, IHD, MI and/or angina pectoris was also significantly associated (GrimAA, OR 1·016 [1·007–1·026], p=0·0101; Grim2AA, OR 1·016 [1·008–1·024], p=0·003). However, non-ASCVD disorders such as chronic kidney disease, cancers, chronic lung disease, asthma, and arthritis were not associated (**Table 4**).

**Table 4.** Age, sex and smoking-adjusted logistic regression analysis of EAA estimated by different epigenetic clocks on cardiovascular and non-cardiovascular diseases (comorbidities).

| Dependent variables | Horvath Age Acceleration |  | Hannum Age Acceleration |  | PhenoAge Acceleration |  | GrimAge Acceleration |  | GrimAge2 Acceleration |  |
| --- | --- | --- | --- | --- | --- | --- | --- | --- | --- | --- |
|  | OR (95% CI) | FDR-adjusted p value | OR (95% CI) | FDR-adjusted p value | OR (95% CI) | FDR-adjusted p value | OR (95% CI) | FDR-adjusted p value | OR (95% CI) | FDR-adjusted p value |
| Hypertension | 1·000 (0·993–1·008) | 0·95 | 1·003 (0·995–1·011) | 0·67 | 1·002 (0·996–1·009) | 0·59 | 1·012 (1·000–1·025) | 0·15 | 1·013 (1·002–1·023) | 0·073 |
| Diabetes mellitus | 0·995 (0·989–1·001) | 0·20 | 1·003 (0·996–1·010) | 0·58 | 1·007 (1·002–1·012) | 0·054 | 1·017 (1·006–1·028) | <0·05 | 1·018 (1·009–1·027) | <0·01 |
| Angina pectoris | 1·002 (0·998–1·006) | 0·58 | 1·005 (1·001–1·010) | 0·11 | 1·004 (1·000–1·007) | 0·12 | 1·011 (1·004–1·019) | <0·05 | 1·010 (1·004–1·017) | <0·05 |
| Atrial fibrillation | 1·001 (0·998–1·003) | 0·74 | 0·999 (0·996–1·002) | 0·58 | 1·001 (0·998–1·003) | 0·74 | 0·996 (0·991–1·000) | 0·17 | 0·997 (0·993–1·001) | 0·22 |
| Stroke | 0·998 (0·994–1·003) | 0·67 | 1·002 (0·997–1·007) | 0·58 | 1·003 (1·000–1·007) | 0·18 | 1·010 (1·002–1·018) | 0·054 | 1·009 (1·003–1·016) | <0·05 |
| IHD | 1·001 (0·997–1·006) | 0·67 | 1·001 (0·997–1·006) | 0·68 | 1·003 (0·999–1·006) | 0·20 | 1·010 (1·003–1·017) | <0·05 | 1·010 (1·004–1·016) | <0·05 |
| MI | 1·002 (1·000–1·004) | 0·11 | 1·001 (0·999–1·004) | 0·35 | 1·002 (1·000–1·004) | 0·12 | 1·005 (1·002–1·009) | <0·05 | 1·004 (1·001–1·007) | <0·05 |
| ASCVD (IHD, MI and stroke)* | 1·000 (0·995–1·006) | 0·98 | 1·005 (0·999–1·011) | 0·22 | 1·007 (1·003–1·012) | <0·05 | 1·016 (1·007–1·026) | <0·05 | 1·016 (1·008–1·024) | <0·01 |
| Chronic kidney disease | 1·000 (0·997–1·004) | 0·85 | 1·001 (0·997–1·005) | 0·74 | 1·003 (1·000–1·006) | 0·15 | 1·006 (1·000–1·012) | 0·12 | 1·006 (1·001–1·011) | 0·072 |
| Cancers | 1·003 (0·999–1·007) | 0·20 | 1·002 (0·998–1·006) | 0·58 | 1·004 (1·001–1·007) | 0·07 | 1·006 (0·999–1·012) | 0·21 | 1·005 (1·000–1·011) | 0·17 |
| Chronic lung disease | 1·001 (0·998–1·004) | 0·58 | 1·000 (0·997–1·004) | 0·85 | 1·000 (0·998–1·002) | 1·00 | 1·007 (1·002–1·012) | 0·052 | 1·005 (1·001–1·009) | 0·073 |
| Asthma | 1·001 (0·997–1·004) | 0·81 | 1·001 (0·997–1·004) | 0·82 | 0·999 (0·997–1·002) | 0·79 | 0·999 (0·994–1·005) | 0·85 | 0·999 (0·994–1·004) | 0·74 |
| Arthritis | 1·003 (0·997–1·009) | 0·57 | 1·006 (0·999–1·014) | 0·18 | 1·004 (0·998–1·009) | 0·29 | 1·008 (0·996–1·019) | 0·32 | 1·009 (1·000–1·018) | 0·16 |

ASCVD, atherosclerotic cardiovascular disease; IHD, ischaemic heart disease; MI, myocardial infarction. Odds ratio (OR) was used to predict the fold change of each dependent variable per unit increase in EAA.
\*Composite variable of IHD, MI and stroke.

### 4. Interrelationship between EAA, systemic inflammation and lipoproteins

Recognising an interrelationship between ASCVD and EAA, we tested the association of EAA and GlycA, a composite biosignature of systemic inflammation by ^1^H-NMR spectroscopy.[Bell 1987; Fung 2023] PhenoAA, GrimAA and Grim2AA were significantly associated with GlycA levels (PhenoAA; adjusted β, 0·005 [0·002– 0·008], p=0·00287; GrimAA, adjusted β, 0·019 [0·013–0·024], p<0·0001; Grim2AA, adjusted β, 0·017 [0·012– 0·022], p<0·0001), whereas HorvathAA and HannumAA were not (**Table 3**). We further performed targeted analysis of 31 lipoprotein features against GrimAA and Grim2AA, and identified 11 and 12 lipid/lipoprotein attributes, respectively, that were significantly associated (**Figure 2**). Specifically, 7 and 8 features associated with GrimAA and Grim2AA, respectively, were related to very-low-density lipoprotein (VLDL), whereas the remaining were largely concordant between GrimAA and Grim2AA — 2 LDL-related (i.e. LDL triglyceride and LDL particle size), 1 HDL-related (i.e. HDL triglyceride and/or HDL particle size), and 1 triglyceride-related (i.e. total triglycerides). Of note, the triglycerides in VLDL, LDL and HDL were consistently associated with GrimAA and Grim2AA (**Figure 2 & Appendix p 7**).

**Figure 2.**
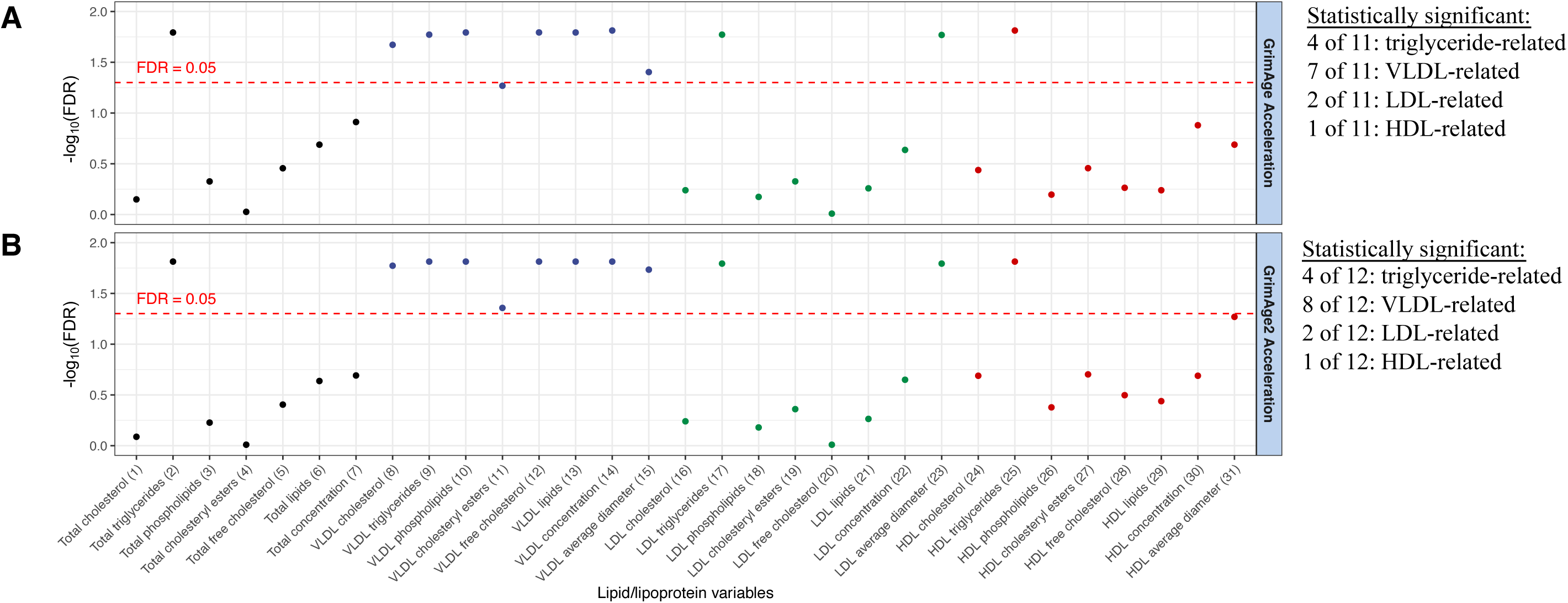
Association analysis of EAA estimated by A) GrimAge or B) GrimAge2 with 31 ^1^H-NMR lipid/lipoprotein features. See Appendix p 7, for list of raw p values.

### 5. LDL particle size is significantly associated with systemic inflammation and frailty

We next tested the association between serum concentration of GlycA and frailty, and found increasing GlycA levels to be significantly higher with incremental frailty (p<0.0001) (**Figure 3**). The median GlycA levels in the robust, pre-frail and frail groups were 1.050 (IQR 0.938–1.197), 1.096 (0.977–1.249) and 1.174 (1.001–1.390) mmol/l, respectively (**Figure 3**). Using partial correlation, we showed that the two GrimAA- and Grim2AA-associated lipoprotein variables, LDL particle size (average diameter) and VLDL particle size (average diameter) (**Figure 2)**, were also associated with GlycA (p<0·001) (**Figure 4)**. Notably, smaller LDL particle size was significantly associated with increased GlycA concentration (*R* = −0·45, p=0·0113) and incremental frailty (robust: 23.83 [23.68–23.90], pre-frail: 23.78 [23.67–23.89], frail: 23.76 [23.66–23.85], Jonckheere-Terpstra trend test, p=0.0145; (**Figure 4B**)), whereas larger VLDL particle size (average diameter) was positively correlated with GlycA concentration (*R* = 0·52, p=0·0357) but inversely related to frailty (robust: 39.42 (38.30–40.58), pre-frail: 39.20 (38.21–40.62), frail: 39.05 (37.81– 40.37) (**Figure 4D**). Other lipid/lipoprotein variables were positively correlated with GlycA concentration but not associated with frailty (**Figure 4 & Appendix p 8**).

**Figure 3.**
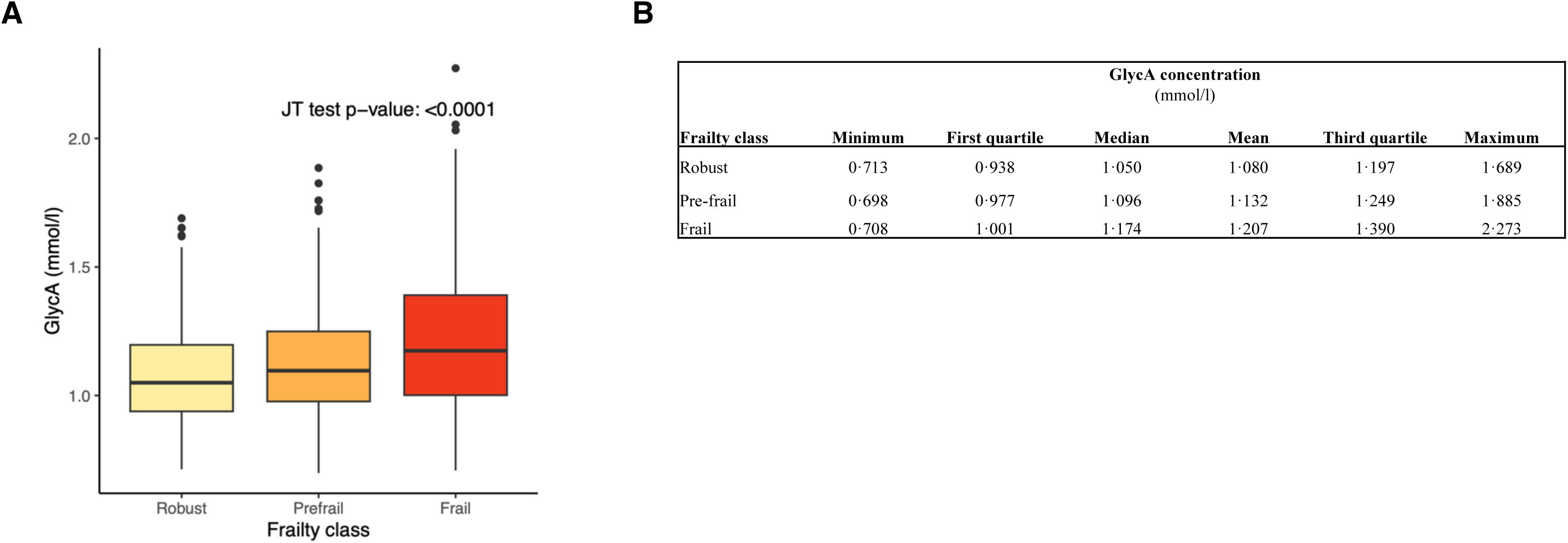
**A**) & **B**) Positive association between frailty and systemic inflammation indicated by glycosylated acetyls (GlycA). The Jonckheere-Terpstra (JT) test was used to calculate the statistical significance.

**Figure 4.**
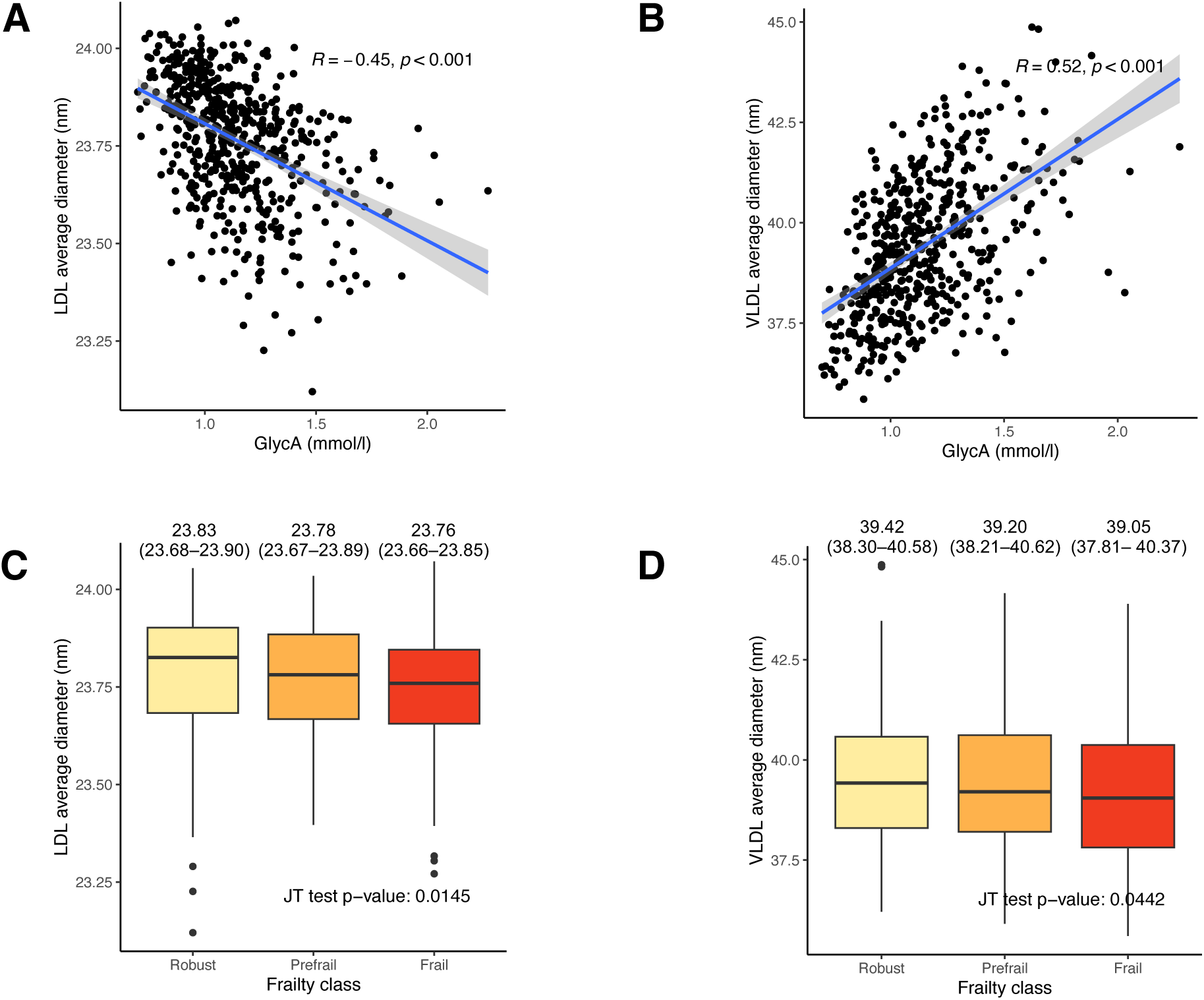
Two of 12 significant findings from Figure 2 with significant association with frailty are shown. **A**) LDL particle size (average diameter, nm) is negatively correlated with GlycA concentration. **B**) VLDL particle size (average diameter, nm) is positively correlated with GlycA concentration. **C**) LDL particle size and **D**) VLDL particle size are inversely associated with frailty class. See Appendix p 8 for the remaining 10 of 12 significant correlations between lipid/lipoprotiens and GlycA but without an association with frailty class. JT test, Jonckheere-Terpstra trend test; LDL, low-density lipoprotein; VLDL, very low-density lipoprotein.

### 6. EAA, ASCVD risk and incident adverse outcomes

In the context of clinically relevant ASCVD risk prediction, we demonstrated using multiple regression adjusted for age, sex, smoking and systolic blood pressure that increased GrimAA and Grim2AA, but not HorvathAA, HannumAA or PhenoAA, were significantly associated with increased 10-year ASCVD risk calculated by the ACC/AHA ASCVD Risk Calculator (Figure 5). We determined that each 10-year increment in GrimAA and GrimAge2 was associated with increases of 4·45% (p=0·0049) and 4·92% (p=0·0002), respectively, in ASCVD risk.

**Figure 5.**
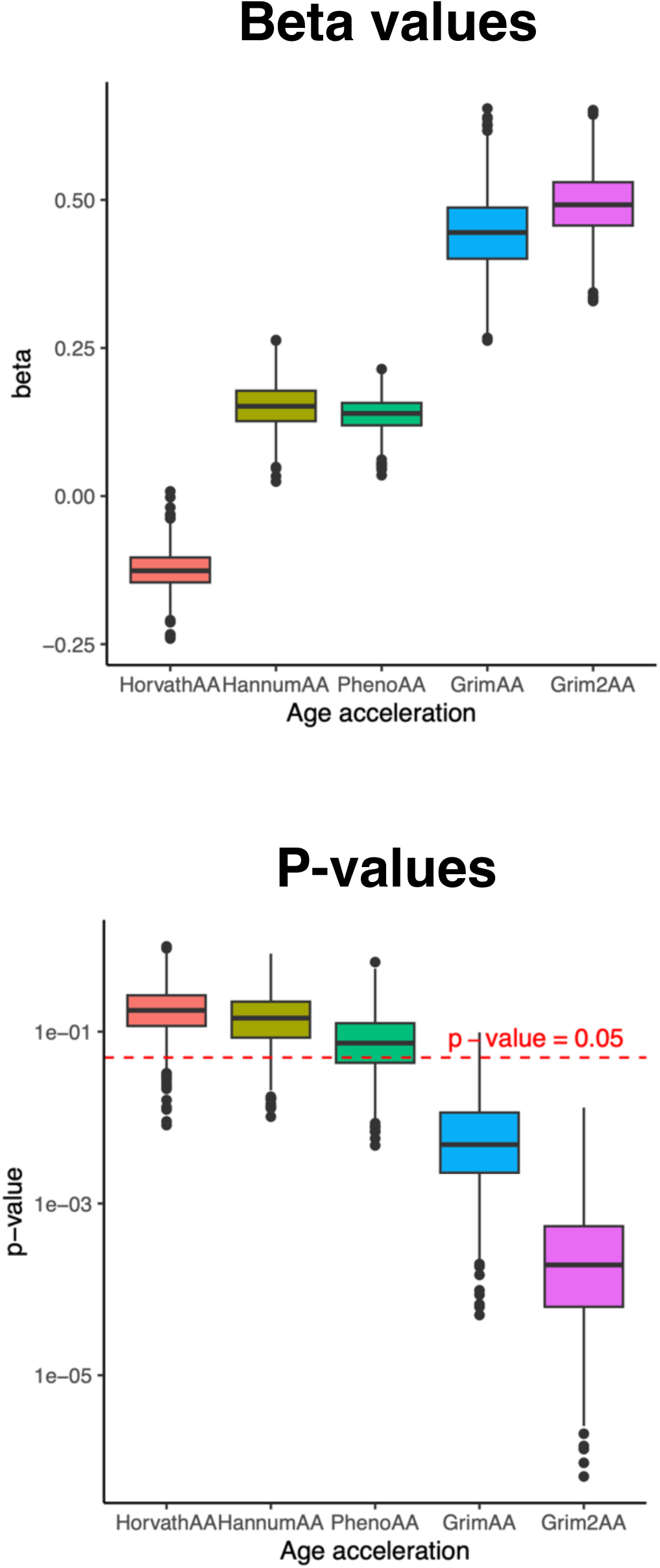
Association analysis of EAA calculated by different epigenetic clocks against 10-year ASCVD risks estimated by the AHA/ACC Risk Calculator.

**Figure 6.**
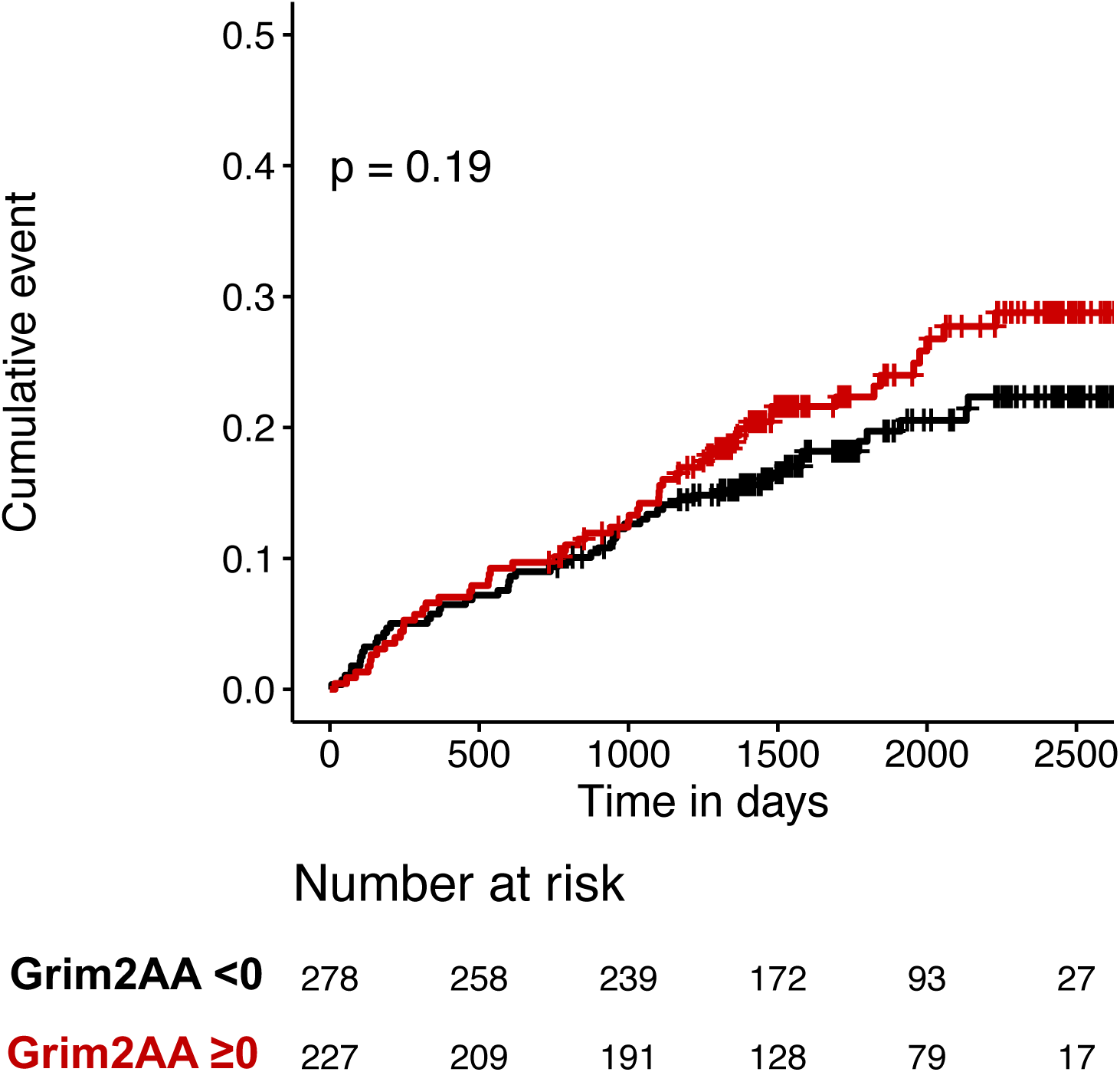
Kaplan-Meier analysis of cumulative events of cardiovascular hospitalisation stratified by Grim2AA <0 or ≥0 years.

In our prospective observational cohort (30 individuals not included due to missing outcomes), 55 of 505 (10·9%) older adults died during a median follow-up of 4·68 years (IQR 3·91–6·44; min 1·85, max 7·14), and 101 (20·0%) were hospitalised for any cardiovascular reason (cardiovascular hospitalisation). In all Cox proportional hazards models tested, chronological age had the largest effect size and strongest level of statistical significance (**Tables 5–7**). Grim2AA was determined to be an independent predictor of incident cardiovascular hospitalisation, whereas GlycA and/or LDL particle size did not have a significant effect (**Table 6**). Among older adults with cardiovascular hospitalisation, the median Grim2AA was 0·112 years (IQR −1·855, 2·898), whereas for those without cardiovascular hospitalisation, it was −0·568 years (IQR −2·981, 2·150). With each 1-year increase in Grim2AA, the risk for cardiovascular hospitalisation was 6·1% (p=0·0162) in Model 1 and 5·7% (p=0·0318) in Model 4 (**Table 6**). However, Kaplan-Meier analysis did not suggest a significant difference in the rate of cardiovascular hospitalisation between patients with Grim2AA <0 and those with Grim2AA ≥0 years.

**Table 5.** Cox proportional hazard models for all-cause mortality (58 of 505 individuals).

|  | **Model 1** |  | **Model 2** |  | **Model 3** |  | **Model 4** |  |
| --- | --- | --- | --- | --- | --- | --- | --- | --- |
| Dependent variables | HR (95% CI) | p value | HR (95% CI) | p value | HR (95% CI) | p value | HR (95% CI) | p value |
| Age (y) | 1·10 (1·06–1·14) | <0·0001 | 1·11 (1·07–1·15) | <0·0001 | 1·10 (1·06–1·14) | <0·0001 | 1·10 (1·06–1·15) | <0·0001 |
| Sex | 1·50 (0·76–2·94) | 0·24 | 1·82 (0·92–3·59) | 0·086 | 1·61 (0·82–3·14) | 0·17 | 1·75 (0·85–3·59) | 0·13 |
| Smoking | 1·59 (0·79–3·18) | 0·19 | 1·68 (0·84–3·38) | 0·15 | 1·78 (0·88–3·58) | 0·11 | 1·60 (0·79–3·24) | 0·20 |
| Grim2AA (y) | 1·04 (0·98–1·11) | 0·20 |  |  |  |  | 1·03 (0·96–1·10) | 0·45 |
| GlycA (mmol/l) |  |  | 2·56 (0·89–7·34) | 0·080 |  |  | 2·75 (0·81–9·30) | 0·10 |
| LDL particle size (nm) |  |  |  |  | 1·167 (0·193–7·042) | 0·87 | 2·69 (0·36–20·21) | 0·34 |

Model 1: Hazard (death) = age + sex + smoking + Grim2AA
Model 2: Hazard (death) = age + sex + smoking + GlycA
Model 3: Hazard (death) = age + sex + smoking + LDL particle size
Model 4: Hazard (death) = age + sex + smoking + Grim2AA + GlycA + LDL particle size

**Table 6.**
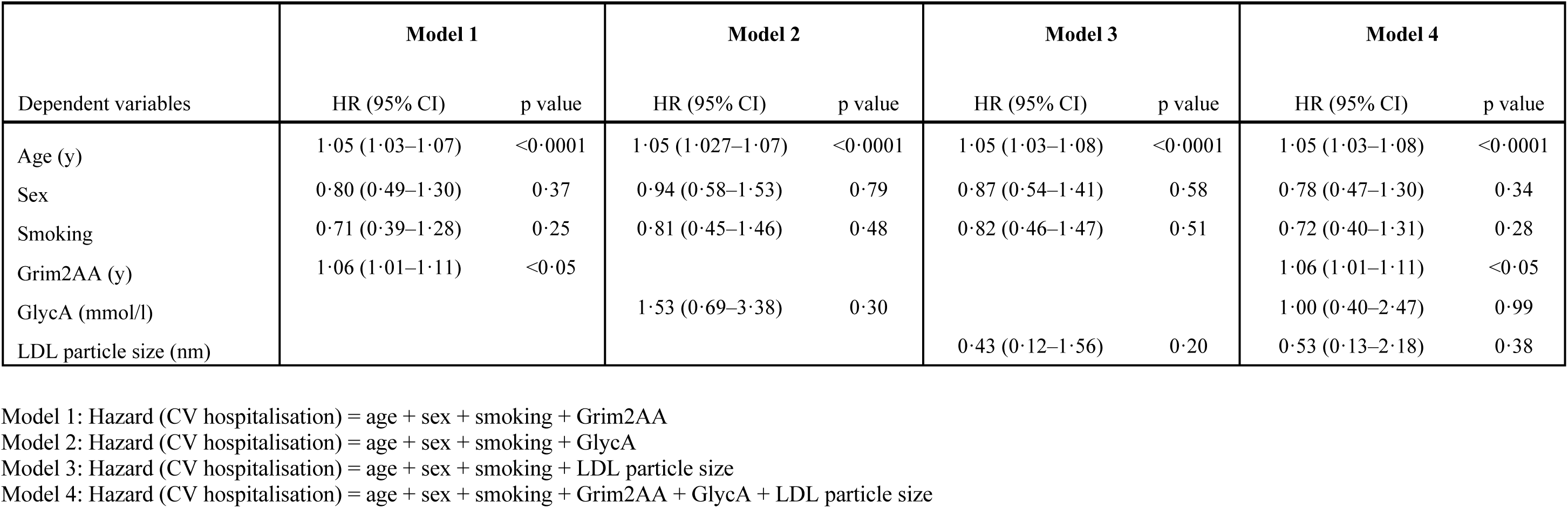
Cox proportional hazards models for cardiovascular hospitalisation (101 of 505 individuals).

**Table 7.** Cox proportional hazards models for ASCVD hospitalisation (42 of 505 individuals).

|  | **Model 1** |  | **Model 2** |  | **Model 3** |  | **Model 4** |  |
| --- | --- | --- | --- | --- | --- | --- | --- | --- |
| Dependent variables | HR (95% CI) | p value | HR (95% CI) | p value | HR (95% CI) | p value | HR (95% CI) | p value |
| Age (y) | 1·08 (1·04–1·12) | <0·001 | 1·08 (1·04–1·13) | <0·0001 | 1·09 (1·04–1·13) | <0·0001 | 1·084 (1·04–1·13) | <0·0001 |
| Sex | 0·68 (0·31–1·49) | 0·33 | 0·97 (0·44–2·15) | 0·94 | 0·83 (0·38–1·80) | 0·64 | 0·67 (0·29–1·54) | 0·34 |
| Smoking | 1·22 (0·53–2·82) | 0·64 | 1·50 (0·64–3·51) | 0·35 | 1·55 (0·67–3·62) | 0·31 | 1·25 (0·54–2·92) | 0·60 |
| Grim2AA (y) | 1·11 (1·04–1·20) | <0·01 |  |  |  |  | 1·10 (1·02–1·20) | <0·05 |
| GlycA (mmol/l) |  |  | 2·67 (0·80–8·96) | 0·11 |  |  | 1·15 (0·27–4·83) | 0·85 |
| LDL particle size (nm) |  |  |  |  | 0·19 (0·03–1·44) | 0·11 | 0·32 (0·03–3·00) | 0·32 |

Model 1: Hazard (ASCVD hospitalisation) = age + sex + smoking + Grim2AA
Model 2: Hazard (ASCVD hospitalisation) = age + sex + smoking + GlycA
Model 3: Hazard (ASCVD hospitalisation) = age + sex + smoking + LDL particle size
Model 4: Hazard (ASCVD hospitalisation) = age + sex + smoking + Grim2AA + GlycA + LDL particle size

Among patients with cardiovascular hospitalisation, ASCVD (including acute coronary syndrome and peripheral vascular disease such as acute limb ischaemia and aortic aneurysm) excluding stroke was the commonest reason for cardiovascular hospitalisation (42 of 101 individuals, 41·6%) with the highest median Grim2AA of 0·73 years (**Figure 7**). Stroke by itself (20 of 101 individuals, 19·8%) was associated with a reduced Grim2AA of −0.85 years. Interestingly, permanent pacemaker implantation (n=5) for complete heart block was associated with the most strongly reduced Grim2AA of −1·43 years. Importantly, Grim2AA ≥0 years was found to be a strongly significant predictor of future hospitalisation due to ASCVD (ASCVD hospitalisation, p=0·0027) during median follow-up of 4·68 years.

**Figure 7.**
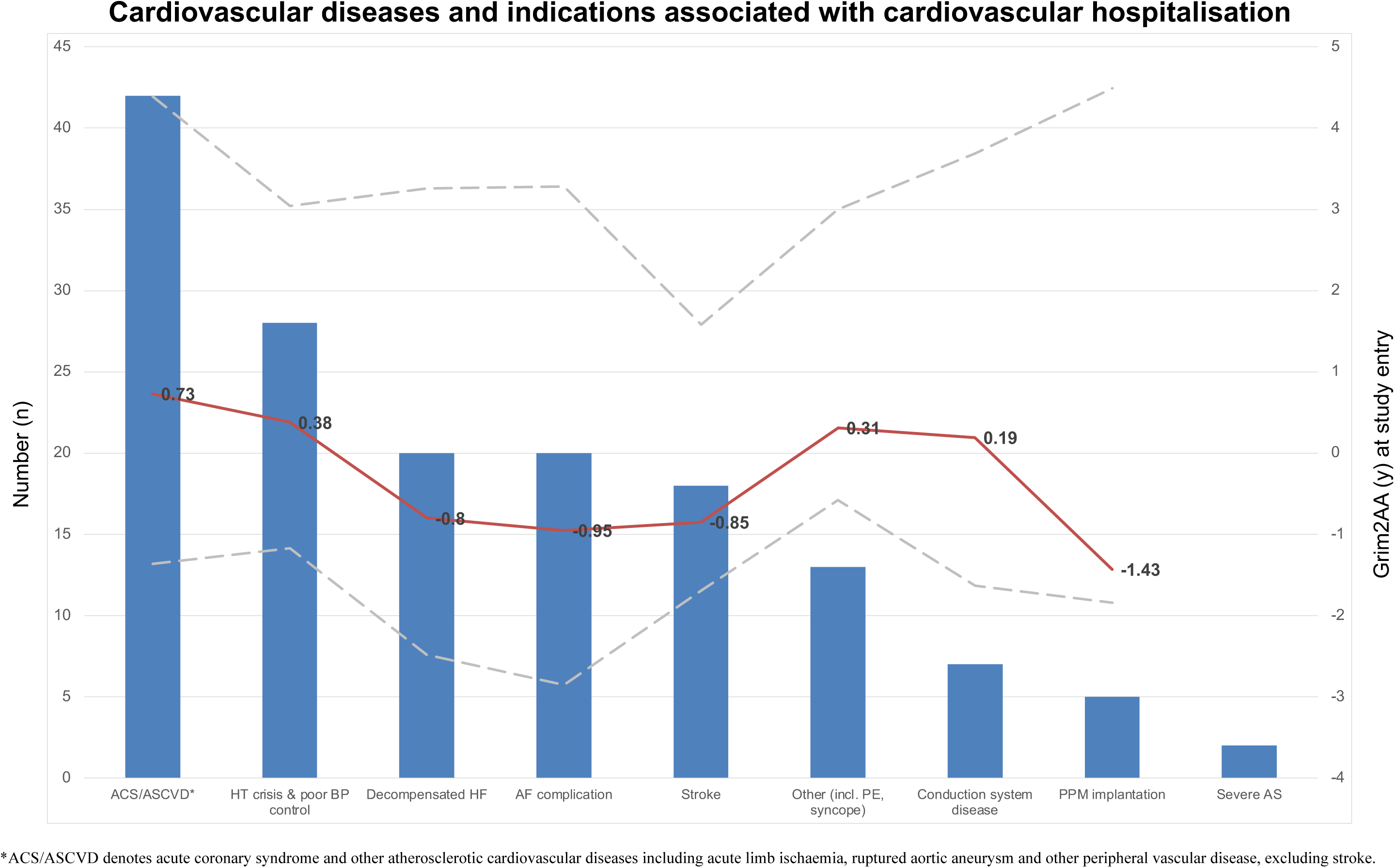
Frequency distribution of cardiovascular diseases and indications (in *blue*) associated with incident cardiovascular hospitalisation in 101 older adults during a median follow-up of 4·68 years (interquartile range (IQR) 3·91–6·44). The median Grim2AA (years; in *red*) and IQR (Q1, Q3; in *grey*) values at study entry are plotted along the secondary y-axis and shown for each condition.

**Figure 8.**
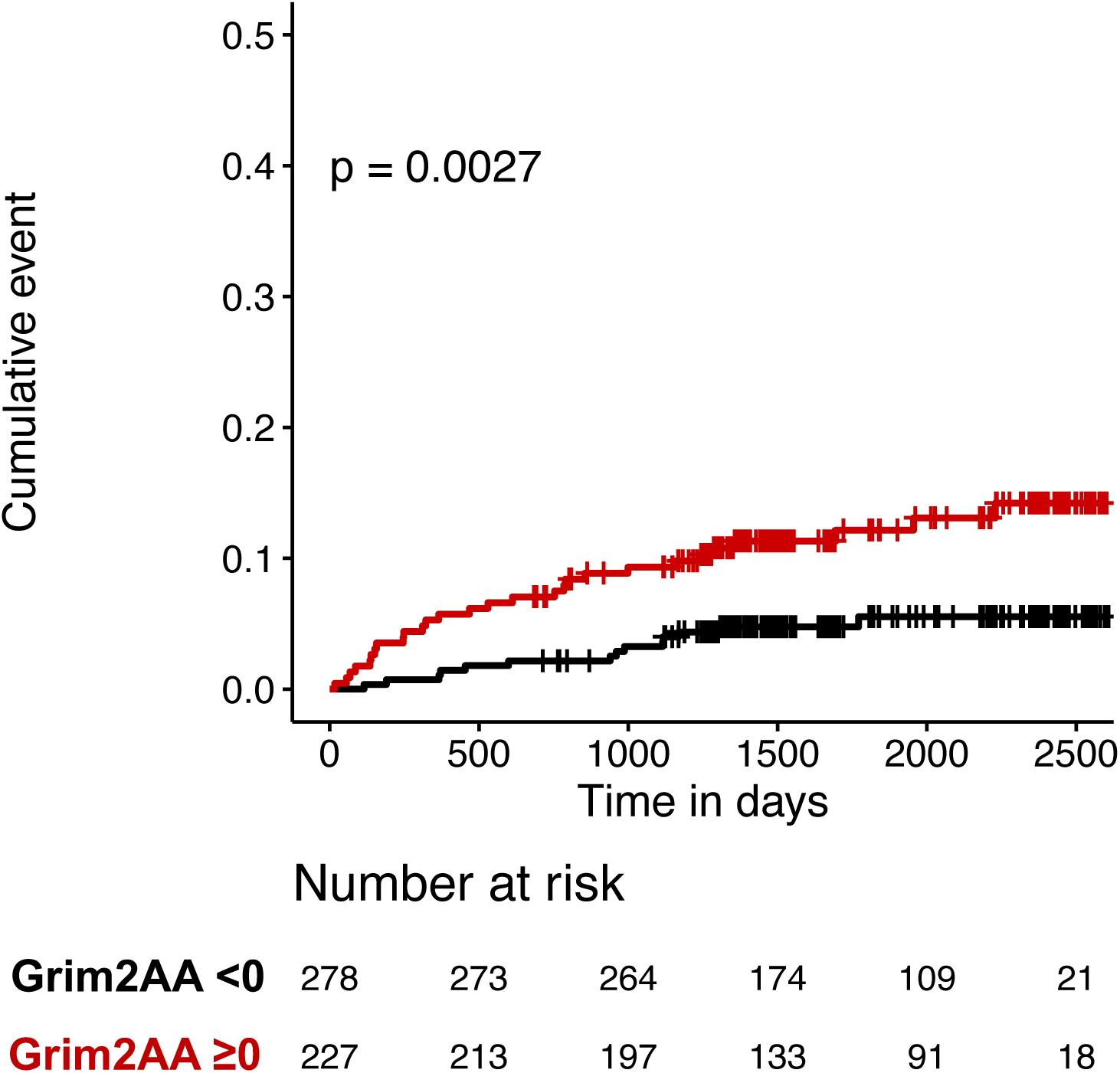
Kaplan-Meier analysis of cumulative events of ASCVD hospitalisation stratified by Grim2AA <0 or ≥0 years.

## Discussion

This study represents the most comprehensive targeted analysis of circulating blood lipid-metabolome and DNA methylation-based epigenetic clock linking frailty and EAA (particularly, GrimAA and Grim2AA) with atherogenic dyslipidaemia, systemic inflammation and ASCVD in older adults. The clinical importance of our findings were explored using the ACC/AHA ASCVD Risk Calculator, and further verified through prospective analysis of incident adverse cardiovascular outcomes in our study cohort. In particular, we identified a prognostic role for Grim2AA as an epigenetic biomarker of ASCVD risks and an independent predictor of future ASCVD hospitalisation precipitated by serious cardiovascular diseases including acute coronary syndrome, acute limb ischaemia, unruptured or ruptured aneurysm, among others. We have also linked increased Grim2AA to the pathophysiology of frailty and atherogenic dyslipidaemia involving LDL particle size and systemic inflammation as reflected in circulating GlycA concentration. Although LDL particle size and GlycA levels were not informative for prediction of future adverse cardiovascular outcomes in older adults, our findings may open avenues for further mechanistic studies on inflammageing.

Previous studies have reported separate associations between EAA and frailty, inflammation or ASCVD risks. But none has detailed the characteristics of atherogenic dyslipidaemia associated with inflammageing and EAA in connection with frailty. The main findings from this community-based cohort of older adults including the old-old and oldest old were: 1) smaller LDL particle size (average diameter) was significantly associated with incremental frailty and increased systemic inflammation; 2) systemic inflammation was associated with increased EAA (particularly, GrimAA and Grim2AA) and frailty; 3) increased VLDL particle size and other VLDL-associated attributes were significantly associated with increased systemic inflammation (Figure 4B) and GrimAA or Grim2AA (Figure 2) but, interestingly, not frailty (Figure 4D); 4) TRLs including VLDL and other lipoprotein-triglycerides were associated with increased GrimAA and Grim2AA (Figure 2); 5) Grim2AA provided the most informative estimate of cardiometabolic risks linking atherogenic dyslipidaemia, inflammageing, frailty, and incident adverse outcomes primarily associated with ASCVD.

The inflammatory pathways and mechanisms underlying atherogenesis and ASCVD associated with ageing have been summarised previously.[Ferrucci 2018; Libby 2021; Aranda 2024; Kraler 2025] However, the pathobiology of vascular inflammation, ageing and frailty remains poorly understood. The role of GlycA as a biomarker of systemic inflammation in cardiometabolic disorders has garnered increased interests owing to its positive correlation with CRP, low threshold of detection in blood, and high intra-assay repeatability (low percent coefficient of variation).[Fung 2023] GlycA is a composite ^1^H-NMR signal from N-acetyl methyl groups of N-acetylglucosamine in blood glycoproteins that increases with inflammation,[Bell 1987] infection (e.g. sepsis), and in cardiometabolic disorders ranging from pre-diabetes, early diabetes mellitus to subclinical atherosclerotic vascular disease.[Ritchie 2015; Fung 2023] Previous epidemiological studies have shown its usefulness in tracking systemic inflammation over time, given its relative stability and association with disease and disease risks. We demonstrated in this study that GlycA reflected existing ASCVD comorbidities, provided a useful correlate for association analysis with atherogenic dyslipidaemia, and differentially indicated inflammation across the frailty spectrum.

In a recent systematic review of 12 studies among which 9 had used the Fried or modified Fried frailty phenotype (e.g. FRAIL scale), the authors summarised that the associations between dyslipidaemia and frailty were inconsistent and unclear.[Shakya 2022] Of note, elevated triglyceride levels were highlighted as being associated with frailty in multiple cross-sectional and longitudinal studies, in contrast to one study reporting lower HDL associated with new-onset frailty.[Shakya 2022] In a recent comprehensive narrative review by James et al., chronic inflammation mediated by proinflammatory cytokines, immune activation, cellular changes, and metabolic dysregulation were pointed out as major abnormal biological changes associated with cardiovascular diseases and frailty; however, the interrelationship and mechanisms underlying the pathophysiology remain unknown.[James 2024] We and others have previously shown that early-life indicators in the lipid-metabolome can inform about cardiometabolic risks in adulthood.[Ojanen 2021] Those biomarkers included LDL particle size (average diameter), VLDL-triglycerides, ratio of apolipoprotein B to apolipoprotein A-1 (ApoB/ApoA1), other lipoprotein features (e.g. HDL particle size, large HDL-phospholipid) and GlycA that indicated increased cardiometabolic risks.[Ojanen 2021; Fung 2023] In our current study of older adults, LDL particle size, VLDL subfractions and particles including VLDL-triglyceride, and GlycA levels were abnormal, and validated some of the previous findings from unrelated study cohorts. A UK Biobank study recently reported that VLDL particle size was significantly associated with coronary heart disease, [Jin 2023] in line with our observation that VLDL particle size was strongly and positively correlated with GlycA. However, the marginally significant inverse association between VLDL particle size and frailty may seem aberrant and suggest a chance association or a complex non-linear interrelationship that is not understood. The non-linear cardiometabolic risks associated with VLDL (very small or extremely large subclasses) have been reported previously.[Jin 2023]

In this study, we consistently found that Grim2AA was most strongly associated with ASCVD risk factors and manifestations, compared with other epigenetic clocks. This may stem from the design of Grim2AA using constituent DMPs in algorithms derived from machine learning-selected surrogate markers, including growth differentiation factor 15 (GDF15), cystatin C, CRP and haemoglobin A1c, that have important pathophysiological functions in, or connection with, inflammageing, cardiometabolic disease and frailty.[Lu 2019] With advances in personalised medicine incorporating multi-omic approaches including DNA methylation and metabolomic information, it is feasible to integrate EAA with the blood metabolome as an alternative or complementary high-dimensional bioanalytic approach to ASCVD risk estimation.

There are several limitations in this study. The study was designed to have robust, pre-frail and frail older adults in similar ratios through selection of study participants from the parent cohort of approximately 1,400 individuals. Because frail older adults are markedly less common in the community than pre-frail and robust individuals, it is therefore obvious that the selection strategy maximises statistical power and balance between groups at the expense of potential selection bias. We have tried to minimise the effects of bias by first selecting all frail older adults in the cohort, followed by random selection of pre-frail and robust older adults using software algorithm in R. Second, in estimating ASCVD risk, we have used the ACC/AHA ASCVD Risk Calculator designed for the American population. Studies have shown that the risk may be overestimated in the Asian population. Furthermore, the ASCVD risk assessment in older adults aged above 79 years is known to be inaccurate. In view of a lack of data in ASCVD Risk Calculator for the age range above 79 years, the risk estimates can only be recalibrated using real-world prospective data. This study attempted to fill that gap by providing a prospective analysis of incident adverse outcomes, and identified a strong prediction capacity of Grim2AA for future ASCVD hospitalisation. Third, most of the study participants recruited into this cohort were ambulatory and community-living. In the frail group, most of the individuals were mild-to-moderately frail, as reflected in the FRAIL scale and eFI (Table 1). Severely frail older adults were thus not well represented, and institutionalised older adults were not included. This study set out to recruit ambulatory, community-living older adults who could complete a range of cardiogeriatric assessments. A separate study focusing on severely frail older adults may therefore provide another level of understanding of the complex morbidity, risks, and outcomes in challenging care settings that are beyond the scope of the present study.

## Conclusions

EAA estimated by the epigenetic clock, GrimAge 2, provides informative cardiometabolic risk assessment in older adults with no previous history of heart failure that aligns with ASCVD Risk Calculator-generated estimates. Increased Grim2AA is associated with atherogenic dyslipidaemia, inflammageing, frailty and adverse cardiovascular outcomes, particularly ASCVD-related hospitalisation.

## Data Availability

All data produced in the present study are available upon reasonable request to the authors.

## Acknowledgments

The authors thank the Research Grants Council for funding support from the General Research Fund (Ref. No. 14116421 awarded to EF).

