## Appendix for "Epigenetic age acceleration links atherogenic dyslipidaemia, inflammageing and frailty with adverse cardiovascular outcomes in older adults (UFO): a lipid-metabolome and epigenetic clock analysis"

**Supplementary Table 1**. Characteristics of participants in the UFO study of epigenetic age, frailty, inflammageing and ASCVD. Raw p values are shown.

| Variable | All | Robust (FRAIL 0) | Pre-frail (FRAIL 1–2) | Frail (FRAIL ≥3) | Raw p value |
|---|---|---|---|---|---|
| N | 535 | 186 | 180 | 169 | - |
| Median age (Q1–Q3), *y* | 78 (69–85) | 71 (66–83) | 80 (70–85) | 82 (75–86) | 2.283028e-11 |
| Median age (range, min–max), *y* | 78 (60–99) | 71 (60–92) | 80 (60–94) | 82 (63–99) | - |
| Sex (% male), *n* | 177 (33·1%) | 90 (48·4%) | 53 (29·4%) | 34 (20·1%) | 1·295e-08 |
| Body mass index, $kg/m^2$ | 24·03 (21·88–26·62) | 23·46 (21·76–25·42) | 24·52 (21·95–27·16) | 24·35 (21·91–27·27) | 0.01524185 |
| Smoking history (%) | 110 (20·6%) | 49 (26·3%) | 33 (18·3%) | 28 (16·6%) | 0·02141654 |
| Past alcohol use (%) | 116 (21·7%) | 50 (26·9%) | 40 (22·2%) | 26 (15·4%) | 0·008835235 |
| eGFR, $mL/min/1{\cdot}73\ m^2$ | 79·23 (63·24–89·71) | 85·51 (74·50–94·12) | 75·78 (61·07–87·84) | 71·13 (51·01–85·52) | 1·300785e-13 |
| **Frailty measurements** | | | | | |
| eFI | 0·11 (0·06–0·19) | 0·06 (0·03–0·08) | 0·14 (0·08–0·19) | 0·22 (0·17–0·28) | 4·665032e-47 |
| 6-minute walk distance, *m* | 326 (259–386) | 380 (340–426) | 320 (248–380) | 258 (189–308) | 6·379045e-40 |
| Gait speed, *m/s* | 0·92 (0·71–1·13) | 1·17 (1·04–1·30) | 0·86 (0·70–1·02) | 0·73 (0·58–0·87) | 5·385984e-36 |
| Handgrip strength, *kg* | 18 (14–23) | 22 (18–27) | 17 (14–22) | 15 (13–18) | 1·239786e-21 |
| **Comorbidities** | | | | | |
| Hypertension | 315 (58·9%) | 72 (38·7%) | 116 (64·4%) | 127 (75·1%) | 2·259505e-12 |
| Diabetes mellitus | 117 (21·9%) | 14 (7·5%) | 44 (24·4%) | 59 (34·9%) | 3·849006e-10 |
| Angina pectoris | 49 (9·2%) | 3 (1·6%) | 12 (6·7%) | 34 (20·1%) | 2·033226e-09 |
| Atrial fibrillation | 18 (3·4%) | 6 (3·2%) | 7 (3·9%) | 8 (4·7%) | 0·1007281 |
| ASCVD (IHD, MI and stroke)* | 91 (17·0%) | 9 (4·8%) | 31 (17·2%) | 51 (30·2%) | 2·50462e-10 |
| IHD | 46 (8·6%) | 6 (3·2%) | 11 (6·1%) | 29 (17·2%) | 3·574255e-06 |
| MI | 10 (1·9%) | 2 (1·1%) | 1 (0·6%) | 7 (4·1%) | 0·03759142 |
| Stroke | 53 (9·9%) | 2 (1·1%) | 20 (11·1%) | 31 (18·3%) | 4·887728e-08 |
| Chronic kidney disease | 31 (5·8%) | 2 (1·1%) | 7 (3·9%) | 22 (13%) | 1·835668e-06 |

**Supplementary Table 2**. Association between frailty and epigenetic age or epigenetic age acceleration (EAA). All raw p values shown.

| **Characteristic** | **All** | **Robust** (FRAIL 0) | **Pre-frail** (FRAIL 1–2) | **Frail** (FRAIL ≥ 3) | **p value*** |
|---|---|---|---|---|---|
| N | 535 (100%) | 186 (34·8%) | 180 (33·6%) | 169 (31·6%) | |
| **Epigenetic clock** | | | | | |
| Horvath's Age (2013), *y* | 72·79 (66·96–79·24) | 69·71 (64·79–75·92) | 74·20 (67·70–79·92) | 74·52 (70·21–79·71) | 3·15E-07 |
| Hannum's Age (2013), *y* | 65·03 (54·97–68·40) | 60·30 (54·97–68·40) | 65·42 (58·81–72·32) | 67·29 (62·00–72·45) | 1·97E-09 |
| PhenoAge (2018), *y* | 65·40 (57·73–72·81) | 59·90 (54·28–69·42) | 67·20 (60·05–73·52) | 68·27 (62·25–74·43) | 3·33E-10 |
| GrimAge (2019), *y* | 73·74 (66·27–79·44) | 68·20 (62·77–77·69) | 74·50 (67·60–79·46) | 76·50 (70·81–81·37) | 1·27E-11 |
| GrimAge2 (2022), *y* | 79·23 (71·57–84·45) | 73·63 (68·08–82·12) | 79·44 (73·39–84·19) | 81·82 (76·14–86·62) | 1·52E-13 |
| **Epigenetic age acceleration** | | | | | |
| Horvath's AA, *y* | 0·04 (-4·08 to 3·77) | -0·09 (-3·82 to 3·71) | 0·09 (-3·78 to 3·85) | -0·29 (-4·58 to 3·65) | 0·442 |
| Hannum's AA, *y* | -0·45 (-3·58 to 2·97) | -0·50 (-3·15 to 2·69) | -0·29 (-3·23 to 2·93) | -0·52 (-3·99 to 3·17) | 0·981 |
| PhenoAA, *y* | -0·24 (-4·02 to 3·70) | 1·01 (-4·28 to 2·47) | 0·40 (-3·35 to 4·62) | -0·24 (-4·27 to 4·47) | 0·241 |
| GrimAA, *y* | -0·42 (-2·56 to 2·12) | -0·52 (-2·46 to 1·82) | -0·52 (-2·65 to 1·71) | -0·35 (-2·65 to 2·56) | 0·492 |
| Grim2AA, *y* | -0·51 (-2·69 to 2·34) | -0·83 (-3·00 to 1·72) | -0·40 (-2·92 to 2·21) | 0·02 (-2·46 to 3·52) | 0·022 |

**Supplementary Figure 1**. Spearman correlation of epigenetic age with chronological age in male study participants (177 of 535). FRAIL (robust, *grey*; pre-frail, *orange*; frail, *red*) and eFI (non-frail/robust, *grey*; mildly frail, *green*; moderate-to-severely frail, *blue*) classification of frailty classes are shown. All p values <0·0001.

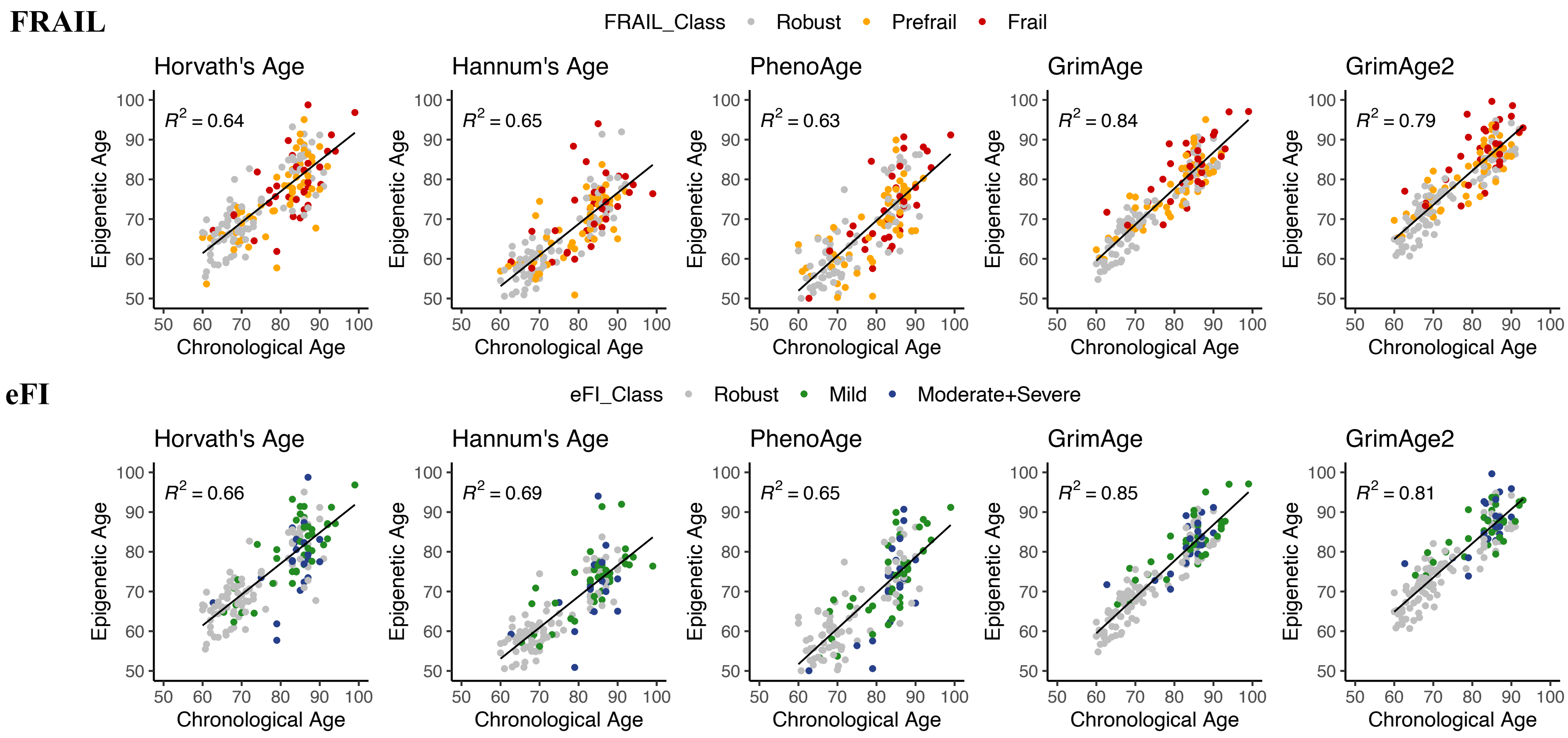

**Supplementary Figure 2**. Spearman correlation of epigenetic age with chronological age in female study participants (358 of 535). FRAIL (robust, *grey*; pre-frail, *orange*; frail, *red*) and eFI (non-frail/robust, *grey*; mildly frail, *green*; moderate-to-severely frail, *blue*) classification of frailty classes are shown. All p values <0·0001.

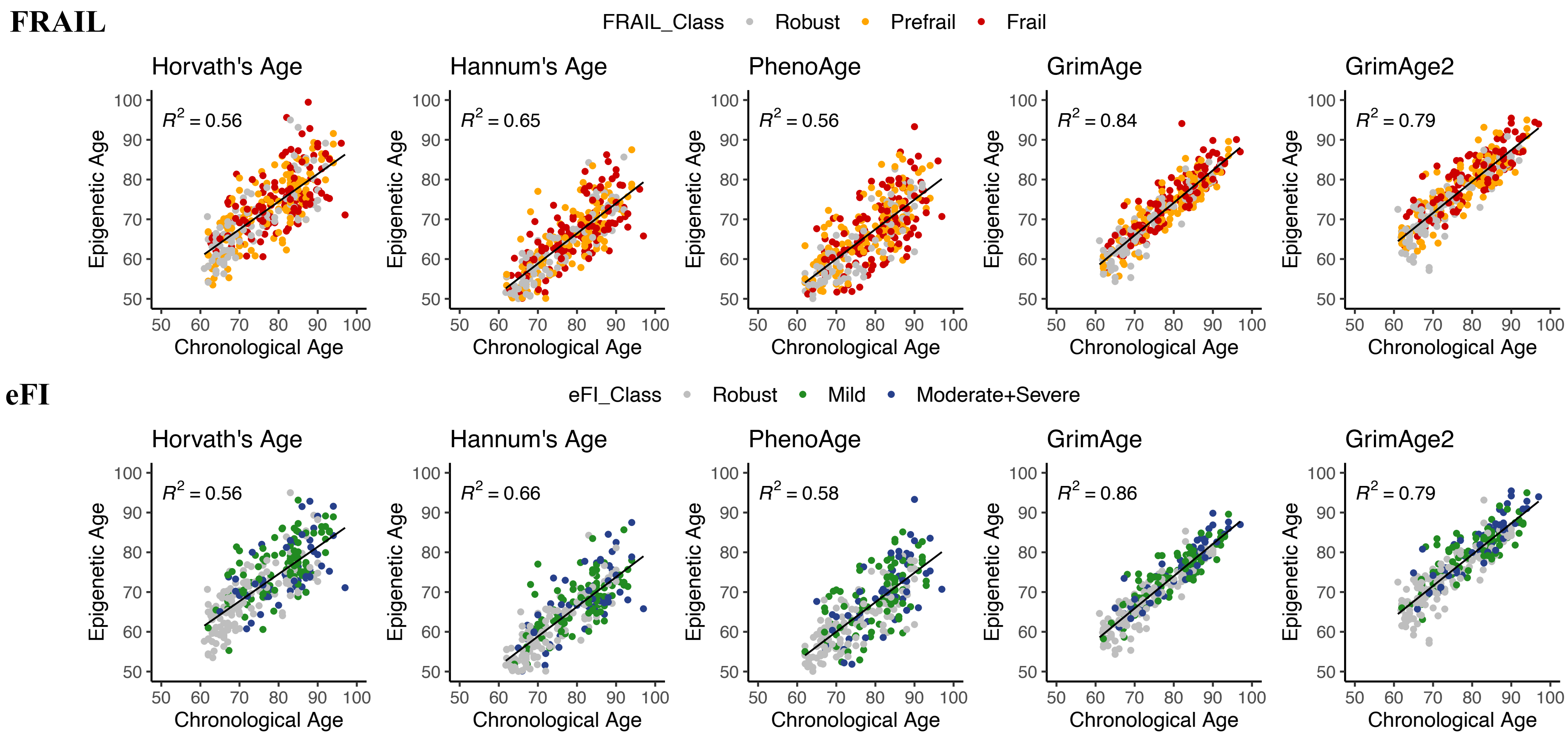

**Supplementary Table 3**. Multiple regression of epigenetic age acceleration (EAA) on FRAIL, eFI, physical fitness measures, and the composite NMR marker of systemic inflammation, GlycA.

| Dependent variables | Horvath Age Acceleration β (95% CI) | Horvath Age Acceleration FDR-adjusted p value | Hannum Age Acceleration β (95% CI) | Hannum Age Acceleration FDR-adjusted p value | PhenoAge Acceleration β (95% CI) | PhenoAge Acceleration FDR-adjusted p value | GrimAge Acceleration β (95% CI) | GrimAge Acceleration FDR-adjusted p value | GrimAge2 Acceleration β (95% CI) | GrimAge2 Acceleration FDR-adjusted p value |
|---|---|---|---|---|---|---|---|---|---|---|
| FRAIL | 0·007 (-0·013 to 0·026) | 0·520345 | 0·02 (-0·001 to 0·041) | 0·091773 | 0·02 (0·004–0·036) | 0·028941 | 0·096 (0·064–0·128) | 4·71E-08 | 0·089 (0·063–0·116) | 1·38E-09 |
| eFI | 0·001 (-0·001 to 0·002) | 0·415385 | 0·002 (0–0·003) | 0·036316 | 0·001 (0–0·002) | 0·061286 | 0·007 (0·004–0·009) | 6·60E-07 | 0·006 (0·004–0·008) | 4·43E-08 |
| 6-min walk distance | -1·177 (-2·538 to 0·184) | 0·117261 | -2·318 (-3·846 to -0·79) | 0·006969 | -1·414 (-2·53 to -0·298) | 0·0262 | -5·336 (-7·633 to -3·039) | 2·29E-05 | -5·022 (-6·92 to -3·125) | 1·25E-06 |
| Gait speed | -0·006 (-0·01 to -0·001) | 0·028941 | -0·009 (-0·014 to -0·004) | 0·00087 | -0·004 (-0·007 to 0) | 0·061286 | -0·017 (-0·024 to -0·01) | 2·29E-05 | -0·014 (-0·02 to -0·008) | 3·51E-05 |
| Handgrip strength | 0·035 (-0·049 to 0·118) | 0·448929 | -0·078 (-0·174 to 0·017) | 0·13375 | -0·01 (-0·08 to 0·06) | 0·773 | -0·168 (-0·309 to -0·027) | 3·32E-02 | -0·155 (-0·272 to -0·038) | 2·09E-02 |
| GlycA | -0·002 (-0·005 to 0·002) | 0·43 | 0·003 (-0·001 to 0·007) | 0·2112 | 0·005 (0·002–0·008) | 0·002153 | 0·019 (0·013–0·024) | 9·43E-09 | 0·017 (0·012–0·022) | 1·77E-10 |

β value indicates the predicted change in the dependent variables per 1-year increase in EAA. eFI, electronic frailty index; FDR, false discovery rate; GlycA, glycosylated acetyls.

**Supplementary Table 4**. Summary of p values from association analysis of EAA estimated by GrimAge and GrimAge2 with 31 1H-NMR lipid/lipoprotein features. Raw p values adjusted for false discovery rate (FDR) are shown.

| No. | Lipid/lipoprotein variables | **GrimAA** p value | **Grim2AA** p value |
|---|---|---|---|
| 1 | Total cholesterol | 0·7092 | 0·8178 |
| 2 | Total triglycerides | 0·0161 | 0·0154 |
| 3 | Total phospholipids | 0·4717 | 0·5931 |
| 4 | Total cholesteryl esters | 0·9405 | 0·9790 |
| 5 | Total free cholesterol | 0·3497 | 0·3941 |
| 6 | Total lipids | 0·2048 | 0·2308 |
| 7 | Total concentration | 0·1226 | 0·2035 |
| 8 | VLDL cholesterol | 0·0213 | 0·0169 |
| 9 | VLDL triglycerides | 0·0169 | 0·0154 |
| 10 | VLDL phospholipids | 0·0161 | 0·0154 |
| 11 | VLDL cholesteryl esters | 0·0538 | 0·0439 |
| 12 | VLDL free cholesterol | 0·0161 | 0·0154 |
| 13 | VLDL lipids | 0·0161 | 0·0154 |
| 14 | VLDL concentration | 0·0154 | 0·0154 |
| 15 | VLDL particle size (average diameter) | 0·0395 | 0·0184 |
| 16 | LDL cholesterol | 0·5759 | 0·5759 |
| 17 | LDL triglycerides | 0·0169 | 0·0161 |
| 18 | LDL phospholipids | 0·6698 | 0·6617 |
| 19 | LDL cholesteryl esters | 0·4712 | 0·4368 |
| 20 | LDL free cholesterol | 0·9790 | 0·9790 |
| 21 | LDL lipids | 0·5517 | 0·5451 |
| 22 | LDL concentration | 0·2308 | 0·2243 |
| 23 | LDL particle size (average diameter) | 0·0170 | 0·0161 |
| 24 | HDL cholesterol | 0·3644 | 0·2048 |
| 25 | HDL triglycerides | 0·0154 | 0·0154 |
| 26 | HDL phospholipids | 0·6361 | 0·4196 |
| 27 | HDL cholesteryl esters | 0·3492 | 0·1986 |
| 28 | HDL free cholesterol | 0·5448 | 0·3184 |
| 29 | HDL lipids | 0·5759 | 0·3644 |
| 30 | HDL concentration | 0·1318 | 0·2048 |
| 31 | HDL particle size (average diameter) | 0·2048 | 0·0538 |

**Supplementary Figure 3**. From Figure 2, the remaining 10 of 12 significant correlations between lipid/lipoprotiens and GlycA but without an association with frailty class are shown: A) total triglyceride, B) VLDL cholesterol, C) VLDL triglycerides, D) VLDL phospholipids, E) VLDL cholesteryl esters, F) VLDL free cholesterol, G) VLDL lipids, H) VLDL concentration, I) LDL triglycerides, and J) HDL triglyerides. The lipid/lipoprotein variable numbers correspond to the ones in Figure 2 and Appendix p 7.

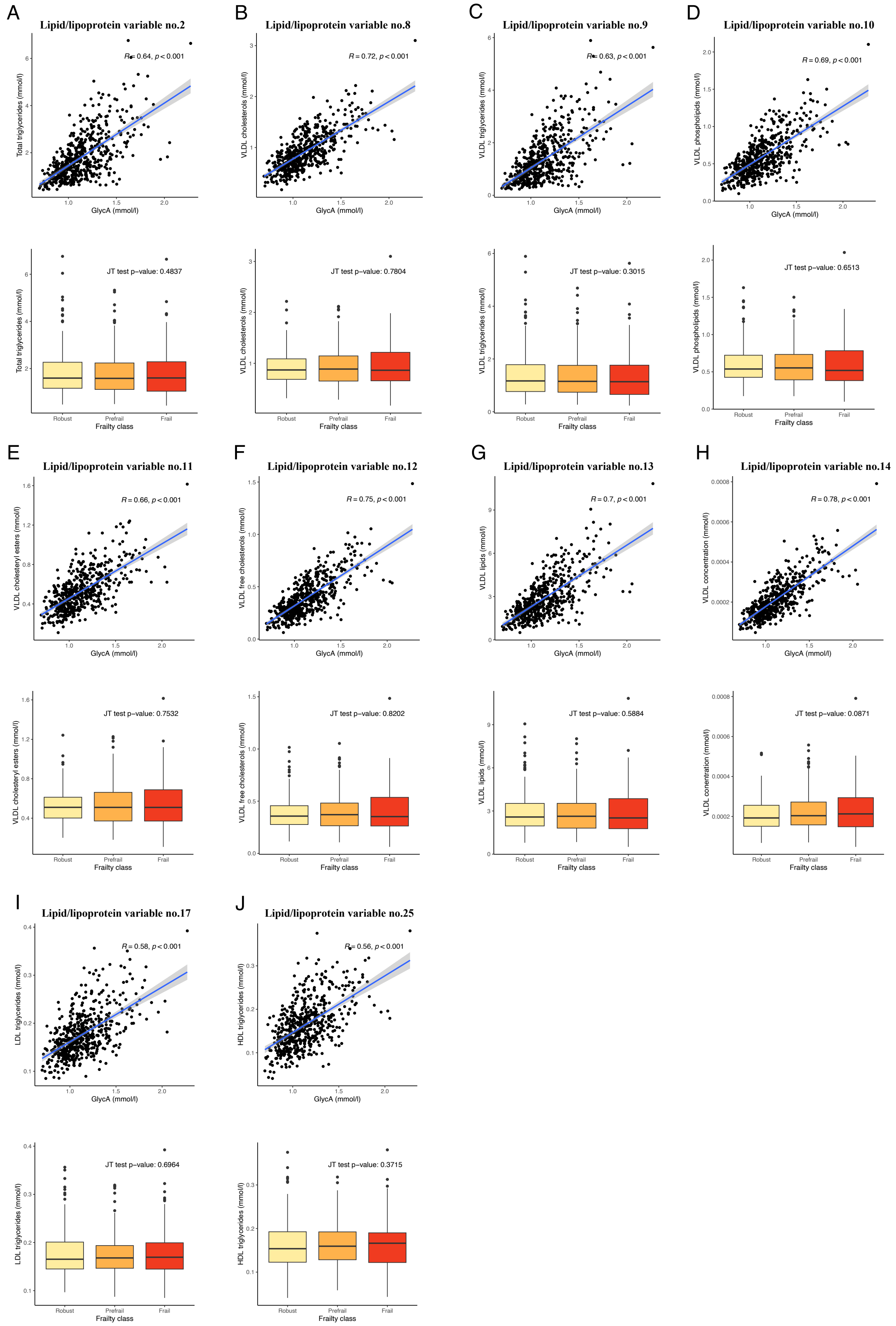
